# Dynamic interactions of influenza viruses in Hong Kong during 1998-2018

**DOI:** 10.1101/19008987

**Authors:** Wan Yang, Eric H. Y. Lau, Benjamin J. Cowling

**Author notes:** Correspondence to: Wan Yang, Department of Epidemiology, Mailman School of Public Health, Columbia University, 722 W 168th Street, Room 514, New York, NY 10032.

## Abstract

Influenza epidemics cause substantial morbidity and mortality every year worldwide. Currently, two influenza A subtypes, A(H1N1) and A(H3N2), and type B viruses co-circulate in humans and infection with one type/subtype could provide cross-protection against the others. However, it remains unclear how such ecologic competition via cross-immunity and antigenic mutations that allow immune escape impact influenza epidemic dynamics at the population level. Here we develop a comprehensive model-inference system and apply it to study the evolutionary and epidemiological dynamics of the three influenza types/subtypes in Hong Kong, a city of global public health significance for influenza epidemic and pandemic control. Utilizing long-term influenza surveillance data since 1998, we are able to estimate the strength of cross-immunity between each virus-pairs, the timing and frequency of punctuated changes in population immunity in response to antigenic mutations in influenza viruses, and key epidemiological parameters over the last 20 years including the 2009 pandemic. We find evidence of cross-immunity in all types/subtypes, with strongest cross-immunity from A(H1N1) against A(H3N2). Our results also suggest that A(H3N2) may undergo antigenic mutations in both summers and winters and thus monitoring the virus in both seasons may be important for vaccine development. Overall, our study reveals intricate epidemiological interactions and underscores the importance of simultaneous monitoring of population immunity, incidence rates, and viral genetic and antigenic changes.

## Introduction

Influenza epidemics recur annually in many regions of the world and cause significant morbidity and mortality, leading to 3 to 5 million severe infections and 291,000 to 646,000 deaths worldwide each year (1, 2). These recurrent epidemics are a combined outcome of viral antigenic changes, interactions among co-circulating influenza viruses, transmission and host immune response. Influenza viruses undergo continual genetic mutations and punctuated antigenic changes, which allow them to escape prior immunity and cause new epidemics (3-6). In addition, currently two influenza A subtypes, A(H1N1) and A(H3N2), and influenza B co-circulate in humans; and infection of a certain type/subtype can result in partial immunity to influenza strains of the same type/subtype as well as strains across subtypes (7-15) and types (16-18). The latter cross-reactive immunity—termed cross-immunity—leads to epidemiological interactions among influenza viruses that, along with viral antigenic changes, shape influenza phylodynamics and epidemic dynamics (19-31). Improving understanding of interactions among influenza viruses and the resulting impact on epidemic dynamics will thus provide insights to aid public health efforts to mitigate the burden of influenza.

As observed epidemic dynamics are the result of the aforementioned key evolutionary and epidemiological processes, in this study, we utilized a long-term influenza incidence surveillance dataset collected in Hong Kong since 1998 to estimate the timing of punctuated antigenic changes, strength of cross-immunity, duration of immunity, as well as other key epidemiological parameters for each of the three types/subtypes. Hong Kong is a subtropical city located in Southeast Asia. Unlike the highly seasonal epidemics in temperate regions, in the subtropics and tropics, influenza epidemics can occur at any time of the year (32, 33) and, often, multiple types/subtypes co-circulate causing co-epidemics of comparable magnitude. The long-term Hong Kong dataset with diverse epidemics thus provides a unique opportunity to study the interplay of different influenza types/subtypes at the population level.

These highly diverse epidemic dynamics, however, also present major challenges for inference on transmission dynamics. In an exploratory analysis, we coupled a commonly used multi-strain susceptible-infected-recovered-susceptible (SIRS) model (24) with a particle filter (34-36), a Bayesian inference method, and applied it to the Hong Kong dataset. While this model-inference system was able to reproduce the observed epidemic pattern, the model state did not appear to converge (see Fig. S1). Three major challenges exist. First, the seasonality of influenza in part shapes the epidemic dynamics and thus should be accounted for in the model. However, unlike the winter-time epidemics in temperate regions that have been well-characterized by seasonal forcing of climate variables (humidity alone or with temperature) (37, 38), to date no models capable of capturing the diverse seasonal pattern in tropical and subtropical climates exist. Second, to estimate epidemiological parameters such as the duration of immunity, the inference system must be able to preserve such long-term epidemic features. Single-pass filters (e.g., the particle filter) did not appear to do so as seen in our exploratory analysis. Multi-pass filters (e.g., iterated filters (39-41)) have such built-in capability, however, are less adept in modeling diverse epidemic dynamics (42). Finally, it is challenging to estimate a large number of parameters needed to characterize the interactions between virus-pairs.

We therefore developed a model-inference system that addresses the above challenges. The basic reproductive number *R*_*0*_ is a key epidemiological parameter describing the transmissibility of an infection, defined as the average number of secondary infections that arise from a primary infection in a naïve population. We thus first estimate, for each influenza type/subtype, the average *R*_*0*_ in each week of the year based on the average annual epidemic cycle, i.e. the empirical seasonal cycle. We then incorporate this *R*_*0*_ seasonal cycle (as the prior distribution) in the model-inference system to control for the seasonality observed in Hong Kong. For the second challenge, we combine the IF2 iterated filtering algorithm (41) with space re-probing (36, 43), a technique that allows continuous exploring of state space. The combined SR-IF2 inference method coupled with the empirical seasonal multi-strain SIRS model is thus able to preserve the long-term epidemic feature while adeptly explore the state space to capture the epidemic dynamics. To estimate parameters in high dimensional state space, we take a multi-round inference strategy; in the first round, we parse the entire state space into subspaces and test each to find the optimal subspace; subsequent rounds then build on the previous one until the model estimates converge. Indeed, testing using model-generated mock epidemics indicates that this model-inference system and optimization strategy is able to recover the true model state variables and parameters (*SI Appendix* and Figs S2 and S3).

Here we apply the validated model-inference system to the Hong Kong dataset and infer how major antigenic evolutions and cross-immunity affect influenza transmission dynamics therein over the last 20 years. Our results reveal intricate ecological interactions among the three co-circulating influenza types/subtypes and their impacts on long-term influenza epidemic dynamics.

## Results

### Overall influenza epidemic characteristics and seasonal cycles in Hong Kong

During Jan 1998—July 2018 (∼20 years), there were 17 A(H1N1), 21 A(H3N2), and 18 B epidemics, respectively (or 13, 19 and 14, respectively, if only counting larger epidemics; see *SI Appendix* and Figs. S4 and S5 for divisions of individual epidemics). Figure 1 shows the superimposed annual cycle for each influenza type/subtype in Hong Kong during the study period and the average cycle excluding 2009 for the A(H1N1) pandemic in that year. All three types/subtypes were able to circulate any time in the year. Epidemics of A(H1N1) and A(H3N2) occurred in both winter and summer with similar magnitudes (Fig 1 A and B). In comparison, for influenza B, major epidemics tended to occur in winter months (Fig 1C). As a result, the estimated *R*_*0*_ seasonal cycle has clear bimodal peaks for the two A subtypes (Fig 1 D and E), although to a less extent for A(H1N1); and for type B, it has only one peak in the winter.

**Fig. 1.**
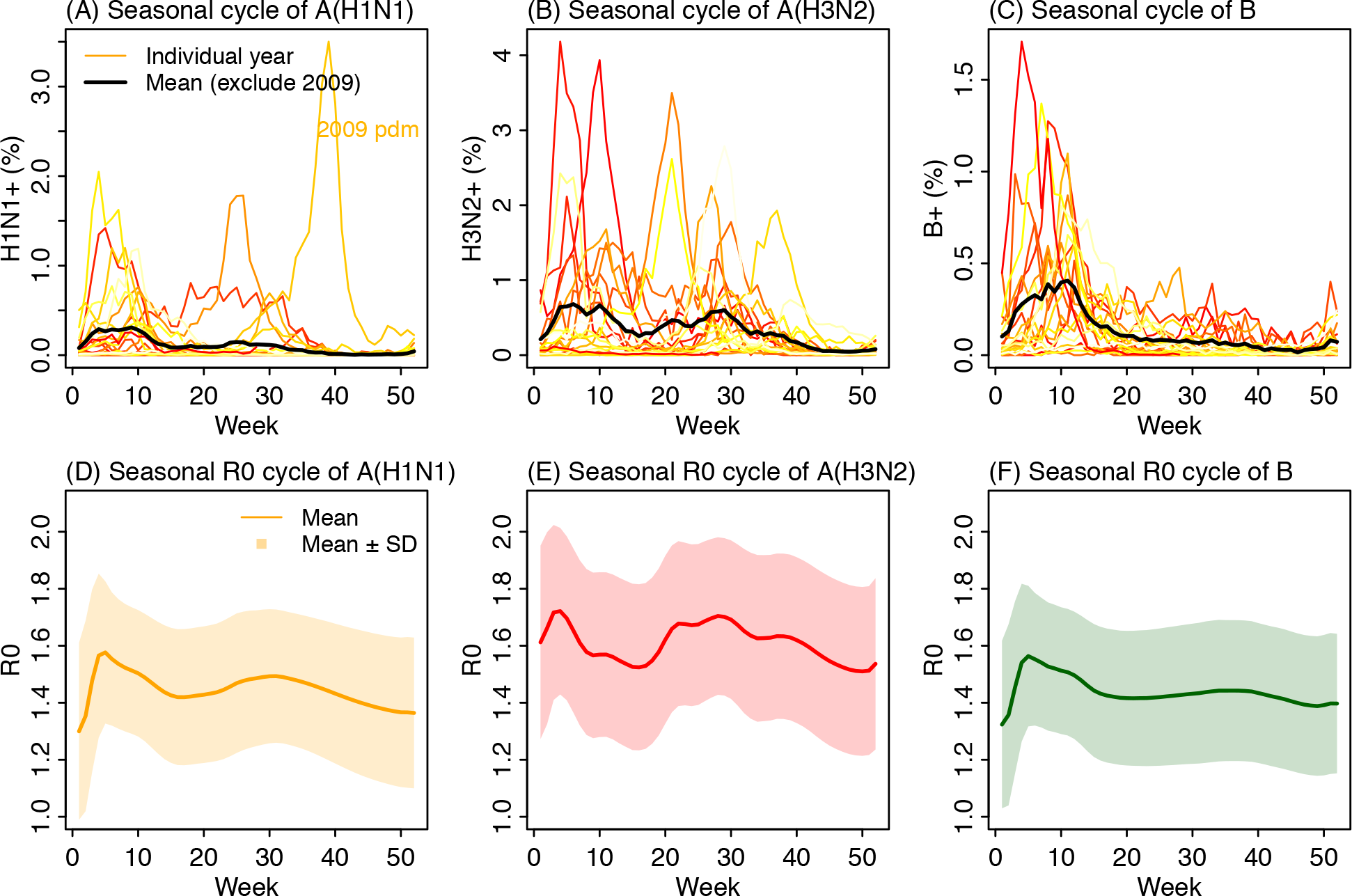
Seasonality of influenza epidemics in Hong Kong. Upper panel: annual cycles for A(H1N1) (A), A(H3N2) (B), and B (C). Color lines show infection rates for individuals years during Jan 1998 –July 2018; lighter colors show earlier years and darker colors for later years. Thick black lines show the mean excluding 2009 for the A(H1N1) pandemic. Lower panel: estimated seasonal *R*_*0*_ cycles for A(H1N1) (D), A(H3N2) (E), and B (F).

### Model-fits to influenza epidemic dynamics in Hong Kong (1998–2018)

Figure 2 shows the model-fits to the epidemic curves of the three influenza types/subtypes. Despite the highly irregular epidemic timing and intensity, the model-inference system, run continuously through the entire 20-year study period, was able to recreate the observed epidemic dynamics for all three types/subtypes. Distributions of residuals were centered at 0, indicating no systematic biases of the model (Fig S6). In addition, the model-inference system was able to accurately estimate the infection rates during both epidemic and non-epidemic periods for all types/subtypes. In particular, over the 20-year study period, epidemics of A(H1N1) were less frequent compared to the other two types/subtypes. For instance, few A(H1N1) cases were reported during Jan 1998 – Dec 1999 (the entire 2-year period) and May 2002 – Dec 2004 (the entire 2.5-year period). While *R*_*0*_ followed the same seasonal cycle as years with epidemics (Fig 3A), the estimated infection rates remained at near-zero over these extended time-periods when A(H1N1) was not in circulation (Fig 2A).

**Fig. 2.**
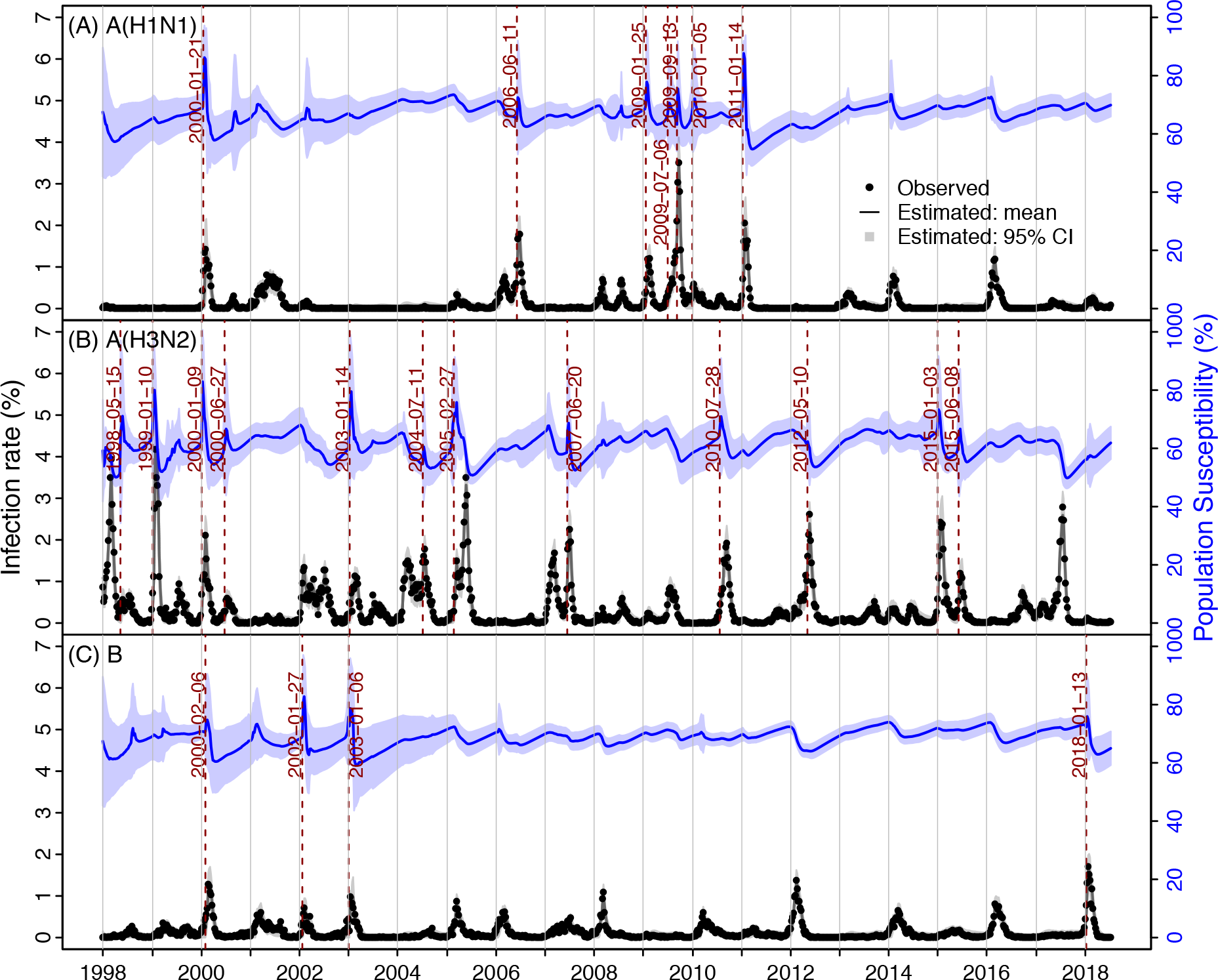
Estimates of weekly infection rate and population susceptibility for the three types/subtypes: A(H1N1) (A), A(H3N2) (B) and B (C). Black dots show observed incidence rates (left y-axis); the lines run through the dots show model estimates; surrounding grey areas show the 95% credible intervals (CI). Blue lines and surrounding areas show the mean and 95% CI estimates of the population susceptibility. Vertical red lines and dates show identified timing of punctuated antigenic changes.

**Fig. 3.**
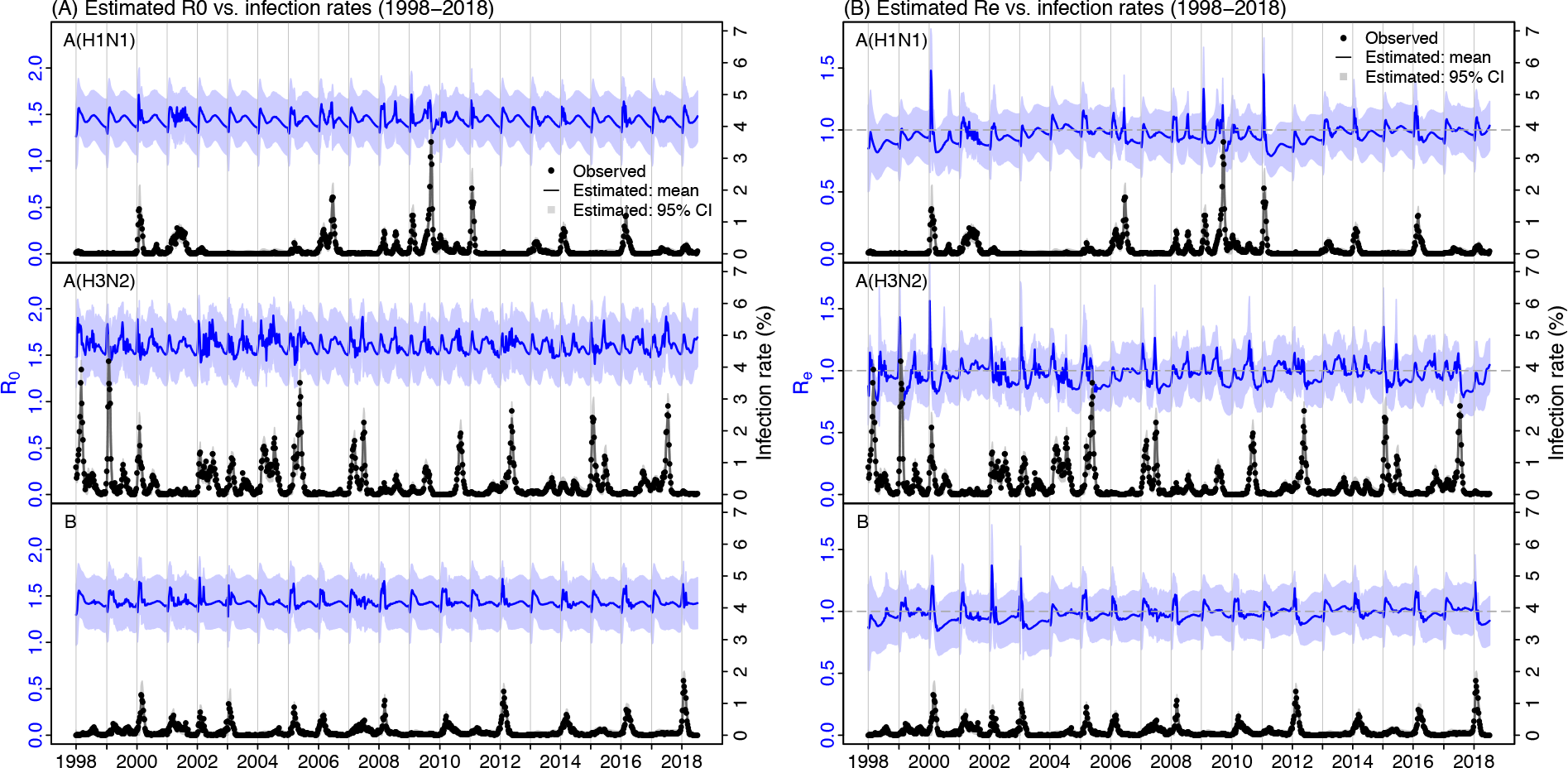
Estimates of weekly reproductive number: basic reproductive number (A) and effective reproductive number (B). Blue lines and areas show the mean and 95% CI estimates. For comparison, the observed (black dots) and estimated (black lines) incidence rates are superimposed (right y-axis).

In addition to running the model continuously without re-initiation to correct for model errors, we included a continuous seeding at a constant rate of 1 case per 100,000 population per 10 days for all types/subtypes. This epidemic seeding represents travel-related importation of cases into local population, since Hong Kong is a transportation hub highly connected to places worldwide. This seeding is also designed to test the accuracy of estimates of key model state variables and parameters, as they are intricately linked to the infection rate. Foremost, overestimation of population susceptibility would lead to overestimation of cases during non-epidemic periods, as the seeded cases would lead to an epidemic should the effective reproductive number *R*_*e*_ (computed as *R*_*0*_ times population susceptibility) be above unity and sustain so. Conversely, underestimation of susceptibility would lead to underestimation of cases during epidemic periods or require unrealistically frequent antigenic changes prior to an epidemic. Further, population susceptibility is also linked to the duration of immunity and the strength of cross-immunity. Loss of immunity (determined by the immunity period) replenishes the susceptible pool whereas gaining of cross-immunity from infection by other types/subtypes (determined by the strength of cross-immunity) can reduce susceptibility. As such, estimates of these related parameters should also be accurate in order to achieve accurate estimation of infection rate. Taken together, the accurate infection rate estimates suggest that the model-inference system was also able to accurately estimate key state variable variables and parameters.

### Population susceptibility, *R*_*0*_, infectious period, and immunity period

Table 1 summarizes the estimates of key epidemiological parameters for each influenza (sub)type. Weekly estimates are shown in Fig 2 (susceptibility), Fig 3 (*R*_*0*_ and *R*_*e*_), and Fig S7 (infectious period, immunity period, and cross-immunity). During Jan 1998 – July 2018, estimated population susceptibility was higher for A(H1N1) and B than A(H3N2) (Fig 2). The mean levels and ranges were 66.6% (range: 50.9–90.3%) for A(H1N1), 60.8% (46.4– 89.0%) for A(H3N2), and 68.6% (50.6–87.8%) for B, respectively. These estimates were consistent with the relative infection rates of individual types/subtypes. Note, however, due to stringent public health interventions (i.e., case isolation, contact tracing, and school closure) implemented from early May to mid-July 2009 (44) and self-protection (e.g., improved personal hygiene and social distancing) during that time (45), the pandemic in Hong Kong unfolded relatively slowly and estimated population susceptibility at the beginning of the 2009 pandemic was relatively low (∼71% in mid-July; Fig 2).

**Table 1.** Estimates of key parameters. Posterior estimates from all runs and weeks are pooled to obtain the mean and full range (in parentheses).

| Strain | Strength of cross-immunity ($c_{ij}$) | | | $R_0$ | Infectious period (days) | Immunity period (years) |
| --- | --- | --- | --- | --- | --- | --- |
|  | A(H1N1) | A(H3N2) | B |  |  |  |
| A(H1N1) |  | 0.17 (0.11, 0.23) | 0.32 (0.26, 0.4) | 1.44 (1.07, 1.82) | 2.64 (2.15, 3.76) | 3.12 (2.64, 3.98) |
| A(H3N2) | 0.4 (0.23, 0.56) |  | 0.24 (0.16, 0.31) | 1.60 (1.32, 1.99) | 3.03 (2.57, 4.30) | 2.28 (1.77, 2.59) |
| B | 0.23 (0.19, 0.32) | 0.11 (0.06, 0.17) |  | 1.43 (1.13, 1.79) | 3.09 (2.15, 4.06) | 3.08 (2.73, 3.48) |

The basic reproductive number *R*_*0*_ was in general higher for A(H3N2) (1.6; range: 1.3–2.0) than A(H1N1) (1.4; 1.1–1.8) and B (1.4; 1.1–1.8). Estimated *R*_*0*_ for the A(H1N1) pandemic in mid-July 2009 was 1.5 (95% CI: 1.3, 1.8), similar to previous estimates (46). The mean effective reproductive number *R*_*e*_, while fluctuated over time (Fig 3B), did average to unity over the 20-year study period for all types/subtypes. The estimated infectious period was slightly longer for A(H3N2) (3.0; 2.6–4.3 days) and B (3.1; 2.2–4.1 days) than A(H1N1) (2.6; 2.2–3.8 days).

The length of immunity period determines the duration of immune protection from prior infection and is thus a key epidemiological parameter shaping the long-term dynamics of influenza epidemics. However, it remains difficult to estimate due to the complex nature of immunogenicity and lack of detail, longitudinal serological data (47, 48). Given the uncertainty, we tested a wide prior range from 1 to 9 years. The posterior estimate of immunity period (Table 1 and Fig S7) was about 3 years for both A(H1N1) (3.1; 2.6–4.0 years) and B (3.1; 2.7–3.5 years), and shorter for A(H3N2) (2.2; 1.8–2.6 years). These estimates accounted for cross-immunity from infections of other types/subtypes, for which, after conversion based on the strength of cross-immunity, was treated the same as specific-immunity from the same type/subtype (see Eqn 1 in Methods).

### Key ecological interactions via cross-immunity

The strength of cross-immunity may modulate the ecological completion among co-circulating influenza viruses. Here we estimated the pair-wise cross-immunity strength, relative to specific-immunity from the same type/subtype (Table 1 and Fig S7; *c*_*ij*_ denotes cross-immunity against virus-*i* gained from infection of virus-*j*). Three interesting features seem to characterize the interactions between the three influenza types/subtypes. First, interactions are asymmetric (i.e., *c*_*ij*_ ≠ *c*_*ji*_) and cross-immunity appears to be weaker from types/subtypes with higher mutation rates. In particular, among all virus-pairs, cross-immunity was the weakest when gained from infection of A(H3N2) (the mean was 17% of the strength of specific-immunity against A(H1N1) and 11% against B). Second, as expected, homotypic-is stronger than heterotypic cross-immunity. Among all virus-pairs, cross-immunity from A(H1N1) against A(H3N2) was the strongest (40%; 23–56%). Third, cross-immunity appears to mediate ecologic competition among the three influenza types/subtypes, with B “controlling” A(H1N1) and the latter further “controlling” A(H3N2). Specifically, the second strongest cross-immunity was from B against A(H1N1) (32%; 26–40%); this, along with the strong cross-immunity from A(H1N1) against A(H3N2), formed a control cascade of B → A(H1N1) → A(H3N2). The higher mutation rate of A(H3N2) virus (4) then allowed it to escape prior immunity including cross-reactive ones from A(H1N1) and B. On the other hand, cross-immunity from A(H1N1) against B (23%; 19–32%) was comparable to the reverse, allowing the two viruses to maintain a more balanced bidirectional ecologic competition.

As our estimates that cross-immunity from A(H1N1) against A(H3N2) is stronger than the reverse appeared to contradict previous notion that A(H3N2) outcompetes A(H1N1) (see, e.g., (49)), we further used model-simulation to test the long-term impact of cross-immunity under three different scenarios: 1) As estimated here, i.e., setting all cross-immunity terms to the posterior mean estimates; 2) A(H3N2) outcompeting A(H1N1) via stronger cross-immunity, for which we set *c*_*H1←H3*_ to 0.5 (vs. 0.4 for *c*_*H3←H1*_); and 3) No interactions, i.e., setting all cross-immunity terms to 0 (see details in *SI Appendix* and simulated epidemics in Figs. S8-10). As shown in Fig. S11, scenario 1 most closely reproduced the observed pattern of co-circulation. In contrast, scenario 2 (i.e., A(H3N2) outcompeting A(H1N1)) largely underestimated the frequency of A(H1N1) epidemics, whereas scenario 3 (i.e., no cross-immunity) largely overestimated the frequency of co-epidemics caused by multiple types/subtypes.

### Timing and frequency of punctuated antigenic changes

Analyses of the antigenic evolution of influenza have shown that major antigenic changes occur in a punctuated fashion, allowing substantial increases in population susceptibility and in turn surge of influenza epidemic (4, 26, 50). To capture such punctuated antigenic changes, our model-inference system allowed abrupt increases in population susceptibility in response to unexpected epidemic surges that are potentially due to major antigenic evolutions in the influenza virus. We then used the major increases in population susceptibility (i.e., >15% increase relative to the preceding week; results using 10% as cutoff are consistent; see Table S1) to identify potential punctuated antigenic changes. Over the 20-year study period, we detected such major changes in population susceptibility 7 times for A(H1N1), including two due to the 2009 pandemic (Fig 2A), 12 times for A(H3N2) (Fig 2B), and 4 times for B (Fig 2C). Table 2 shows the estimated timing of each potential antigenic innovation. Based on these estimates, punctuated antigenic changes occurred every 4.6 (SD=4.6; exponential distribution) years in A(H1N1) (excluding the 2009 pandemic), 1.8 (SD=1.2; gamma distribution) years in A(H3N2), and 6.2 (SD=6.2; exponential distribution) years in B during Jan 1998 – July 2018. These results are consistent with reported estimates based on antigenic testing of influenza virus isolates (4, 50). Further, interestingly, for A(H3N2), such potential antigenic innovations occurred with comparable frequencies in both summers (7 times in May, June or July) and winters (5 times in Jan or Feb) during the 20-year study period. In contrast, it occurred mostly in winters for A(H1N1) and only in winters for B.

## Discussion

Cross-immunity among different influenza viruses has been reported and its resulting potential interactions have been modeled; however, to date, its strength and impact on influenza transmission dynamics at the population level remains unclear. Utilizing a long-term incidence surveillance dataset in Hong Kong since 1998, here we have quantified the strength of cross-immunity among the two influenza A subtypes and B currently co-circulating in humans and the pattern of viral interactions. We have also estimated the timing and frequency of punctuated surges in population susceptibility representing population-level response to antigenic innovations in influenza viruses over the last 20 years as well as identified key variations among three influenza types/subtypes. These estimates, along with other key epidemiological variable estimates including the population susceptibility and basic reproductive number, provide great insight into the evolutionary and epidemiological dynamics of influenza.

### Comparing transmissibility of influenza in Hong Kong and elsewhere

Influenza epidemics in subtropical Hong Kong are highly diverse and frequent (Fig1 and Figs. S4 and S5). Nevertheless, *R*_*0*_ estimates across all three types/subtypes are in the range of 1.1 to 2.0, similar to previous estimates in other regions for influenza overall (51). Few studies have estimated *R*_*0*_ for individual influenza types/subtypes, especially for inter-pandemic seasons. Here we estimate that *R*_*0*_ was similar for A(H1N1) and B (range: 1.1–1.8 for both) and was slightly higher for A(H3N2) (range: 1.3–2.0). Estimated simultaneously with *R*_*0*_, the infectious period, or generation time, was slightly longer for A(H3N2) and B than A(H1N1); these estimates were very close to those based on viral shedding curves—3.0 vs 3.1 days reported for A(H3N2), 3.1 vs 3.4 days reported for B, and 2.6 vs 2.3 days reported for A(H1N1) (52). Together, the higher transmissibility of A(H3N2) (i.e., larger *R*_*0*_ and longer infectious period) in part explains its larger seasonal epidemics and resulting lower average population susceptibility (mean: 60.8% for A(H3N2) vs. 66.6% for A(H1N1) and 68.6% for B). Further, these consistent estimates indicate that our strategy using the empirical *R*_*0*_ seasonal cycle in the model-inference system is effective in controlling for the elusive influenza seasonality in Hong Kong and allows for estimation of other key epidemiological features. Future studies could apply the same strategy to study the diverse influenza dynamics in other subtropical/tropical regions or other infections with similar challenges.

### Cross-immunity and co-circulation pattern

Multiple lines of evidence have suggested cross protection offered by infection of influenza viruses of the same type/subtype as well as across types/subtypes (7-18). However, some studies have reported weak or no heterosubtypic and/or heterotypic cross-immunity (47, 53). As noted by authors of those studies, the cross-sectional serological surveys used therein may miss the window of cross-immunity and/or immunity mechanisms not related to antibodies against the hemagglutinin surface protein (e.g., from T cells mediated immunity). In this study, based on the long-term type/subtype-specific incidence data, we estimate that there were relatively strong cross-immunity against A(H3N2) by infection of A(H1N1) (∼40% of the specific-immunity) and moderate level of cross-immunity between A(H1N1) and B (20-30%). Consistently, our simulations showed that interactions among the three types/subtypes are needed to produce the pattern of co-circulation observed in Hong Kong (Fig. S11 and Fig. S8 vs. Fig. S10).

However, our estimates of weak cross-immunities to A(H1N1) and B conferred by infection of A(H3N2) appear to contradict the common notion that A(H3N2) outcompetes A(H1N1) and B and, as a result, causes more frequent and larger epidemics. For instance, Goldstein et al. (49) showed a negative correlation between early A(H3N2) incidence and subsequent non-A(H3N2) incidence in the same season, suggesting A(H3N2) may interfere with the other two types/subtypes. To test this discrepancy, we simulated epidemics under both scenarios and showed that combining the frequent and large epidemics of A(H3N2) with stronger cross-immunity conferred would likely drive A(H1N1) to extinction (see Fig. S9; note epidemics of A(H1N1) were only possible after the imposed antigenic innovations). Conversely, the strong cross-immunity from A(H1N1) against A(H3N2), as estimated in this study, is consistent with the predominant circulation of A(H1N1) during the 2009 pandemic despite the frequent circulation of A(H3N2) during inter-pandemic seasons.

Thus, to understand the co-circulation pattern of influenza viruses, we need to take into account other key epidemiological features. In particular, A(H3N2) confers shorter immunity following infection and also undergoes more frequent genetic/antigenic mutations. Based on the estimated mean immunity period and implicit assumption of exponential distribution in the model (Eqn 1), the mean half-life of immunity (i.e. ln2 times of the mean) was 1.6 years for A(H3N2), 2.2 years for A(H1N1), and 2.1 years for B. Note our estimates were shorter than reported elsewhere (e.g., 3.5 to 7 years in (47, 48)), likely because, unlike those studies based on titers of antibodies against specific strains, our estimates broadly applied to the entire type/subtype. Nevertheless, the relative lengths of immunity period for the three influenza types/subtypes were consistent. A recent cohort-specific analysis (54) further showed that birth-cohorts whose first exposure was A(H1N1) had long-term protection against subsequent infection by the same subtype whereas those first exposed to A(H3N2) did not (See Table 4 in ref (54)). It is thus likely that A(H3N2) virus’ shorter/weaker immunity and more frequent mutations lead to more frequent A(H3N2) epidemics, and that combining its larger epidemic size and weaker cross-immunity (roughly, the product of these two factors is the decrease in susceptibility to its complement types/subtypes) allows the two less frequently circulating influenza types/subtypes to coexist in humans.

### Estimating timing and frequency of antigenic innovations based on incidence data

Unlike previous analyses of genomic and/or antigenic data, here we tentatively estimate the timing and frequency of punctuated antigenic innovations based on incidence data alone. The rationale is that punctuated antigenic innovations in influenza viruses would lead to surge in population susceptibility and subsequently surge in incidence. That is, incidence data encapsulate the population response to antigenic innovations and thus may complement analyses of genomic and/or antigenic data (note the latter do not provide information as to if an antigenic mutation would lead to an epidemic). Indeed, our results are consistent with estimates from phylogenetic analyses. For instance, the two surges in susceptibility to A(H1N1) during the 2009 pandemic (i.e., around 9/13/09 and 1/5/10; Table 2) roughly matched with the estimated timings of amino acid mutations from NextStrain (55): 1) HA1: S185T and HA2: S124N around 10/1/09 (CI: 7/26/09-1/16/10) and 2) HA1: D97N round 10/30/09 (CI: 09/03/09-02/07/10). In addition, previous studies estimated that punctuated antigenic innovations occur roughly every 2 to 8 years in A(H3N2) (26, 50), 4 to 9 years in A(H1N1), and 3 to 14 years in B (4), based on antigenic analysis of influenza virus isolates. Consistently, we estimate that antigenic innovations occurred very 4.6±4.6 years in A(H1N1), every 1.8±1.2 years in A(H3N2), and every 6.2±6.2 years in B during Jan 1998–July 2018. These consistent results suggest incidence surveillance data could be an inexpensive alternative to support detection of evolutionary changes in influenza viruses, especially since these estimates reflect changes in population immunity in response to antigenic innovations and thus the potential of epidemic surge.

**Table 2.**
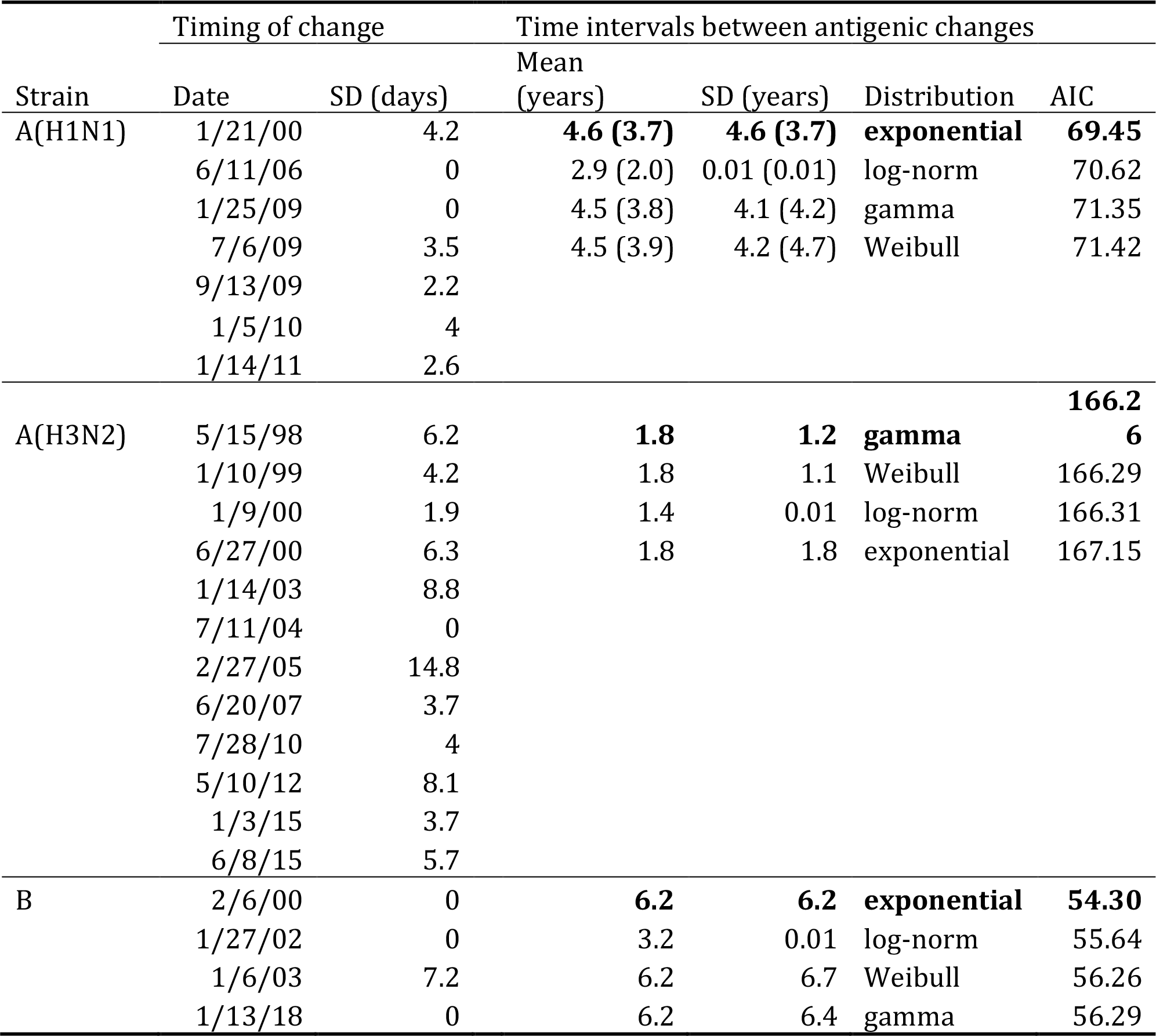
Estimated timing and frequency of potential punctuated antigenic changes. Dates (mm/dd/yy) of potential punctuated antigenic changes are identified based on above 15% increases in susceptibility; the standard deviations are computed based on estimates from all model-inference runs indicating punctuated antigenic changes around the same time. The time intervals between subsequent antigenic changes are estimated based on four different distributions; the best-fit estimates are shown in bold. For A(H1N1), numbers in the parentheses show results when dates during the pandemic are included.

In addition, we note the different seasonal timing of antigenic innovations for A(H3N2). While the identified antigenic innovations predominantly occurred in winters for A(H1N1) and B, we find that A(H3N2) could undergo major antigenic changes in both summers and winters. Further investigation is warranted, as, if confirmed, this seasonality suggests that sampling A(H3N2) strains in the summer may be essential for selection of vaccine strains.

Our study has several limitations. First, as B lineage data were only available since 2014 in Hong Kong, in this study, we did not differentiate the two B lineages (i.e. Yamagata and Victoria). Future work will extend the model-inference system to include and test the epidemiological interactions with the two B lineages. Second, while using the empirical seasonal cycle to account for the epidemic seasonality in Hong Kong has proven critical in this study and may be applied to other subtropical and tropical regions with similar diverse epidemics, better delineation of influenza seasonality in these climates is needed. Third, we assumed that the probability of seeking medical attention for influenza is comparable to other illnesses and, for simplicity, we did not consider the differences in health seeking behavior among patients infected by different influenza (sub)types. Based on this assumption, the estimated annual attack rates ranged from 13% in 2013 to 42% in 2005. There is large uncertainty in estimated influenza attack rates. Recent estimates ranged from 3-11% *symptomatic* infections among vaccinated and unvaccinated US residents during 2010-2016 (56)to 32% confirmed infection among unvaccinated individuals during 2015 in New Zealand (57). In comparison, our estimates for Hong Kong are relatively high but plausible given the longer epidemic duration and lower vaccination rates in Hong Kong (∼10% (58, 59) v. ∼45% in the US (60)). It could also be due to overestimation of A(H3N2) incidence rates, as patients infected with A(H3N2) may be more likely to seek treatment due to more severe symptoms compared to A(H1N1) and B. Further, while we calibrated the incidence rates of A(H1N1) during the 2009 pandemic to the estimated attack rate from serological data (see Methods), our model-inference system did not include public health interventions implemented particularly at the beginning of the pandemic. As a result, our model-inference system—entirely based on incidence data—likely underestimated the increase in susceptibility at the onset of the pandemic. Nevertheless, the estimated timing of antigenic innovations (6 July 2019) was very close to the observed onset of the pandemic in Hong Kong (30 June 2019 (61)). Finally, due to a lack of age-specific incidence data and for simplicity, we did not include age structure in our model. Recent studies have reported differences in immune imprinting in children compared to adults (47, 48). Future work may investigate such age differences further should age-specific incidence data become available.

In summary, using a comprehensive model-inference system, we have revealed in great detail the epidemiological interactions among the three influenza types/subtypes, the antigenic innovations for each type/subtype, and their combined impact on the transmission dynamics of influenza over the last two decades in Hong Kong, a city of global public health significance for influenza epidemic and pandemic control. The intricate epidemiological interactions among influenza viruses and key underlying population features underscore the importance of monitoring population immunity to different influenza viruses, incidence rates, and viral genetic and antigenic changes. Meanwhile, the model-inference system developed here has proven powerful for estimating key evolutionary and epidemiological characteristics of influenza viruses based on type/subtype-specific incidence data and could be extended to study influenza epidemic dynamics in other regions with limited antigenic data.

## Materials and Methods

### Influenza surveillance data

The Centre for Health Protection in Hong Kong monitors seasonal influenza activity through a sentinel surveillance network of approximately 50 private-sector outpatient clinics that report weekly proportion of outpatients presenting with influenza-like illness (ILI; defined as fever >38.0°C plus cough and/or sore throat) (62-64). In addition, the Public Health Laboratory Services Branch thereof conducts laboratory testing for influenza viruses in samples submitted mainly from local hospitals and also a small number of samples submitted by the sentinel outpatient physicians. We obtained weekly records of ILI consultation rate and concurrent (sub)type-specific influenza detection rate from the week ending 04 Jan 1998 to the week ending 14 July 2018. To obtain a more specific measure of the incidence of influenza virus infections in the community, we multiplied the weekly ILI rate by the concurrent viral isolation rate for A(H1N1), A(H3N2) and B, respectively (42, 49, 65, 66). We refer to these combined measures as A(H1N1)+, A(H3N2)+, and B+.

#### Converting ILI consultation rate to infection rate

The ILI consultation rate is the ratio of patients presenting ILI to all patient-visits and does not measure true influenza incidence rate in the catchment population. Previously, we showed that conversion to *per-capita* infection rate can be made based on Bayes’ rule (65, 67). Briefly, ILI+ (here, ILI+ = A(H1N1)+, A(H3N2)+, or B+) estimates the probability that a person seeking medical attention, *m*, has influenza, i.e., *p*(*i*|*m*). By Bayes’ rule, the incidence rate, or probability of a person contracting influenza during a given week, *p*(*i*), is:

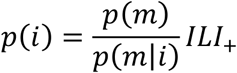

This formula can thus convert ILI+ to the incidence rate *p*(*i*) if 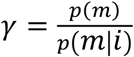 is known. In our previous work, we estimated the conversion factor *γ* along with other model state variables and parameters using a model-inference system without strain-interactions (36). Here to reduce model uncertainty and complexity, we estimated *γ* separately based on independent estimates of attack rate during the pandemic. Based on serological data, estimated attack rates for the 2009 pandemic ranged from 16% (68) to ∼20% (61, 69). Using the upper end of these estimates and the cumulative pandemic H1N1+ rate over the same time period (i.e., 42%), the conversion factor *γ* is ∼0.5 for the pandemic. For the inter-pandemic period, we set *γ*=1, which resulted in annual attack rates ranging from 13% in 2013 and 42% in 2005. The attack rate in 2005 was the highest due to epidemics of all three types/subtypes in the same year (3.3%, 31.1%, and 7.4% attack rates for A(H1N1), A(H3N2), and B, respectively). These estimates are higher than temperate regions but plausible given the year-round circulation of influenza and low vaccination rates in Hong Kong.

### Estimating the empirical seasonal cycle of influenza epidemic

As discussed in the Introduction, the seasonality of influenza epidemics in Hong Kong is weaker than in temperate locations. To model this seasonality, we first computed the weekly average of incidence rate over the study period, excluding year 2009 during which seasonality may be altered due to the A(H1N1) pandemic. We then estimated the time-varying *R*_*0*_ for each week of the year using a model-inference system we previously developed for Hong Kong (36). This system used a basic Susceptible-Infectious-Recovered (SIR) model and ran it with a particle filter with space re-probing. We further smoothed the posterior estimates of *R*_*0*_ using 3-week moving average and used these estimates as the prior in the multi-strain SIRS model-inference system (described below) to represent the empirical seasonal cycle.

### Multi-strain SIRS model

A number of mathematical models have been developed (19-29, 70-73) to study the interplay of influenza viruses. The central idea behind these models is that cross-immunity can either reduce the chance of reinfection and transmission in any individual with prior infection by a similar strain (i.e. leaky immunity) or protect a portion of individuals with prior infection (i.e. polarized immunity). In particular, Gog and Grenfell (24) developed a parsimonious status based multi-strain model by assuming polarized immunity and cross-immunity acted to reduce transmission risk. This model is able to generate evolutionary dynamics similar to that observed for influenza and is commonly used as the backbone for more complex phylodynamic models (26, 28, 73, 74). The multi-strain SIRS model in this study took a construct similar to the Gog & Grenfell model:

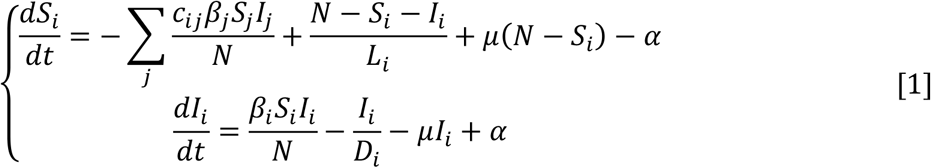

where *N* is the population size; *S*_*i*_ and *I*_*i*_, are, respectively, the numbers susceptible and infected, to virus-*i* (here A(H1N1), A(H3N2), or B); *β*_*i*_, *D*_*i*_, and *L*_*i*_ are, respectively, the transmission rate, mean infectious period, and mean immunity period, for virus-*i*; and *c*_*ij*_ measures the strength of cross-immunity to virus-*i* conferred by infection of virus-*j* (e.g., close to 0 if it is weak and *c*_*ii*_=1 for strain-specific immunity). We assumed the total population size is stable over the study period such that the birth rate and death rate are equal and both are represented by parameter *μ*. The parameter *α* represents travel-related importation of cases and for simplicity was set to 0.1 per 100,000 per day throughout the study period. The model was run continuously in daily time-step from 1 Jan 1998 to 14 July 2018 in conjunction with the SR-IF2 filter (described below), during which variables/parameters were updated weekly at the arrival of observations.

### Inference on transmission dynamics using the SR-IF2 algorithm

We applied a modified IF2 algorithm (termed SR-IF2) to estimate the model state variables and parameters for each week in the 20-year study period. The original IF2 algorithm, developed by and described in detail in Ionides et al (41, 75), implements an iterated, perturbed Bayes map for inference of partially observed dynamic systems. In each iteration, it uses a particle filter (34) to estimate the likelihood of the observed time series (here the A(H1N1)+, A(H3N2)+, and B+ time series altogether). The particle filter uses a suite of model replicates, each is initially randomly drawn from the prior, to represent samples of the latent dynamic variables. Over the course of filtering, the particles are numerically integrated forward in time per the dynamic model (here the multi-strain model in Eqn 1) and resampled based on the likelihood to update the distribution of all model parameters/variables per Bayes’ rule at each time step (here each week). At the end of the time series, model parameters are recycled as starting parameters for the next iteration. In theory, this procedure converges to the maximum likelihood estimate.

While the original IF2 incorporates perturbations of model parameters, our preliminary tests showed that these perturbations alone are insufficient to capture the unexpected surges in population susceptibility due to potential punctuated antigenic changes in the influenza virus. To tackle this issue, we further incorporated space re-probing (SR) in the IF2. Briefly, the SR step allows a small fraction (here 1%) of the model ensemble to sample from the full prior range for key model variables/parameters (here, the susceptibility), whenever filter divergence is detected (i.e., the likelihood drops below a threshold); in addition, to capture other possible changes in parameters related to the virus (e.g., *R*_*0*_ and strength cross-immunity), we set time points requiring SR as break-points, for which larger perturbations are applied to all model parameters/variables (75).

The priors for initiating the SR-IF2 were uniform distributions as follows: *S*_*i*_∼U[50%, 90%] of total population and *D*_*i*_∼U[2, 4] days for *i*=A(H1N1), A(H3N2), or B. To account for seasonality of each (sub)type, the prior of *R*_*0*_ for week *t* of the year was drawn from a uniform distribution with a range of the mean empirical estimate for that week plus/minus one standard deviation. Correspondingly, the prior of transmission rate *β*_*i*_ for week *t* of the year was then computed as *R*_*0,i*_(*t*)/*D*_*i*_(*t*). For the immunity period *L*_*i*_, given the wide range reported in the literature (from months to ∼8 years (76, 77)), we tested prior ranges from 1 to 9 years, divided into four 2-year segments (i.e., [1, 3], [3, 5], [5, 7], and [7, 9] years) for each type/subtype; this resulted in 4^3^=64 combinations for the three type/subtypes. Cross-immunity has been observed for influenza strains of the same subtype as well as across types/subtypes (7-15) and types (16-18); however, the strength has rarely been reported. We thus tested prior ranges from 0% to 80% of the strength of specific immunity, divided into two levels: [0%, 40%] (low) and [40%, 80%] (high); this resulted in 2^6^=64 combinations for 6 virus-pairs (*c*_*ij*_, *i*=H1N1, H3N2, or B and *ji*). To reduce the number of combinations, *a priori*, we further assumed that heterotypic cross-immunity is low especially when the immunity period (*L*_*i*_) is long; thus, for *L*_*i*_>5 years, we only included combinations with low A(H1N1) (or H3N2) and B interactions. Note, however, with perturbations in the IF2 procedure, the posterior estimates can go outside the prior range. Together, we tested a total of 736 combinations of *L*_*i*_ and *c*_*ij*_ priors in the first round of search. We then selected the top ∼1% runs with the best model-fits (highest likelihood *and* fewest space re-probing needed) used their posterior estimates to compute the prior range for subsequent round of search. We repeated this process until the parameters converge. It took two rounds for the synthetic data and three rounds for the observed data.

For each SR-IF2 model-inference run, we performed 50 iterations of filtering with 5000 particles. Each run took round 8 to 10 hours to complete on a high-performance cluster (https://www.mailman.columbia.edu/information-for/information-technology/high-performance-computing-hpc). To account for stochasticity in model initiation, we performed 5 runs for each prior in the first round and 50 runs in the second and third round (i.e., final round for the observed data). From the final round, we selected ∼10 runs with the best model-fits (highest likelihood *and* fewest space re-probing needed) and computed the posterior estimates across those runs per Rubin’s rule (78, 79).

### Estimating the timing and frequency of punctuated antigenic change

To identify potential punctuated antigenic changes for each (sub)type, we computed the changes in susceptibility for each week and considered weeks with a relative *increase* of 15% due to punctuated antigenic changes. We also tested a cutoff of 10%, for which the identified weeks were the same except for 2 additional weeks for A(H3N2) (1/10/1998 and 6/13/1999) and 1 for B (1/28/2001); these additional weeks were all in the early phase of the study period and thus likely due to greater uncertainty in the model-inference system.

To estimate the mean time interval between punctuated antigenic changes, we computed the time intervals between subsequent antigenic changes (right censored for the last observation) and fitted these to four survival models (i.e., the exponential, Weibull, gamma, and log-normal distribution) using the ‘flexsurv’ package (80) in R (https://www.r-project.org). We selected the best model estimate based on the Akaike Information Criterion (AIC).

## Data Availability

The influenza surveillance data are publically available from the Center for Health Protection: https://www.chp.gov.hk/en/static/24015.html; https://www.chp.gov.hk/en/statistics/data/10/641/642/2274.html; https://www.chp.gov.hk/en/statistics/data/10/641/643/2275.html.

## Acknowledgements

We thank the Hong Kong Center for Health Protection for providing data on sentinel surveillance and laboratory surveillance for influenza. We also thank Columbia University Mailman School of Public Health for access to high performance computing.

## SI Appendix

### 1. Validation of the model-inference system

#### 1.1 Method

We first tested our multi-strain SIRS model-SR-IF2 inference system using model-generated mock epidemics (i.e. synthetic data). Specifically, we tested its ability to estimate three main epidemic features of interest: 1) The population susceptibility for each virus over time, including timing of major susceptibility increase due to epochal antigenic change. For the latter, we tested three levels of magnitude—small, medium, and large change—with 10%, 15%, and 25% increase in susceptibility, respectively. 2) The strength of cross-immunity between each virus-pair. For this, we tested two combinations of heterosubtypic immunity (i.e., low H3N2→H1N1 and high H1N1→H3N2 cross-immunity for the first synthetic dataset; high H3N2→H1N1 and median H1N1→H3N2 cross-immunity for the second). and 3) The duration of immunity. Reported immunity period for influenza varied substantially, ranging from months to ∼8 years (1-4). Thus, we tested prior ranges from 1 to 9 years.

To generate the mock epidemics for the above testing, we first generated 100,000 random combinations of initial conditions and model parameters except *R*_*0*_, using Latin Hypercube sampling. Weekly *R*_*0*_ values were randomly sampled from a normal distribution with the mean set to the empirical estimate for that week (Fig 1 in the main text) and standard deviation set to 5% of the mean. To mimic punctuated antigenic changes, for each (sub)type, we first identified the observed major epidemic peak weeks (peak ILI+ in the upper ∼75%, 60%, and 75% percentiles for the three (sub)types, respectively); for each selected peak week, we increased the susceptibility at the 8^th^ preceding week (i.e., roughly the onset) by 15% for A(H1N1) (except for the pandemic) and A(H3N2), by 10% for B, and by 25% for the A(H1N1) pandemic (see the selected weeks in Figs S2 and S3). We then ran the multi-strain SIRS model (Eqn 1 in the main text) for each setting and selected 2 simulations that most closely resembled the observed epidemics in Hong Kong but had different levels of cross-immunity between the two A subtypes: the first simulated time series had high cross-immunity from A(H1N1) against A(H3N2) but low cross-immunity in the reverse direction; and the second had moderate cross-immunity from A(H1N1) against A(H3N2) and high cross-immunity from A(H3N2) against A(H1N1). We then added Poisson random noise to the two simulated time series to mimic observational errors and used these as mock data (termed synthetic ‘truth’) to test the model-inference system.

For each of the two synthetic truths, we performed two rounds of model-inference using the same procedure and prior distributions for the Hong Kong dataset as described in the main text. To evaluate the accuracy of the final posterior estimates, we computed the root-mean-square-error and correlation with the ‘truth’.

#### 1.2 Results

Figures S2 and S3 show the posterior estimates of key model state variables (population susceptibility for the three types/subtypes) and parameters (the basic reproductive number *R*_*0*_, infectious period, immunity period and strength of cross-immunity) for the two tests, respectively, compared with the true values. The relative root-mean-square-error was 0.16 and the correlation was 0.83 combining all model variables/parameters from both tests. These results suggest that our model-inference system and parameter estimation strategy is able to accurately estimate the model state variables and parameters. However, due to large fluctuations of *R*_*0*_ in the synthetic datasets (e.g., Fig S2B) and the collinearity between *R*_*0*_ and susceptibility, the filter tended to compensate sudden increases in infections (due to increases in susceptibility to mimic punctuated antigenic changes) with increases in *R*_*0*_ rather than susceptibility. As a result, it failed to identify some of the punctuated antigenic changes, in particular, for the two A subtypes with larger variances of *R*_*0*_.

### 2. Simulating the impact of cross-immunity on long-term epidemic pattern

#### 2.1 Method

To test the impact of cross-immunity, we simulated epidemics under three different scenarios: 1) As estimated in this study, by setting all cross-immunity terms to the posterior mean estimates; 2) Stronger cross-immunity from A(H3N2) against A(H1N1), by setting *c*_*H1←H3*_ to 0.5, i.e., slightly above the mean estimate for *c*_*H3←H1*_ (i.e. 0.4); and 3) No interactions, by setting all cross-immunity terms to 0. For each simulation, we initiated the multi-strain SIRS model (Eqn 1 in the main text) using the posterior estimates at Week 4 (i.e., after discarding the first three weeks of filter spin-off) and ran the model stochastically from Jan 1998 to July 2018. Weekly values of model parameters (i.e., the infectious period, immunity period, *R*_*0*_, and cross-immunity terms) not specified in the above scenarios were set to the mean posterior estimates from the Hong Kong dataset. Weekly susceptibilities to the three types/subtypes were simulated by the model according to model-simulated infections, loss of immunity, cross-immunity, replenishment from newborns, and deaths; however, to simulate the impact from antigenic innovations, we reset the susceptibility to the posterior mean estimate from the Hong Kong dataset for weeks identified to experience punctuated antigenic changes (Table 1). As in the model-inference runs, we set the epidemic seeding (*α* in Eqn 1) to 0.1 per 100,000 per day for the all simulations. To account for model stochasticity, we simulated each scenario for 1000 times.

To compute the number of epidemics over the study period, as in (5), we defined the epidemic baseline, for each type/subtype, as the 40% quantile of the non-zero corresponding ILI+ records over the ∼20 year study period. We then identified 1) all epidemics including small ones, defined as periods with ≥3 consecutive weeks with ILI+ above the baseline and at least one of the weeks with ILI+ ≥3 times of the baseline; and 2) all large epidemics, defined as periods with ≥6 consecutive weeks with ILI+ above the baseline and at least one of the weeks with ILI+ ≥6 times of the baseline. If an identified epidemic lasted for >1 year, we divided it into multiple epidemics at the year division(s). Based on the 1^st^ definition, there were 17 A(H1N1), 21 A(H3N2), and 18 B epidemics, respectively (Fig. S4); and based on the 2^nd^ definition, there were 13, 19, and 14 epidemics of the three types/subtypes, respectively, during Jan 1998—July 2018 (Fig. S5). We used the same baselines as observed and definitions to identify and compute the numbers of epidemics in the simulations and compared them to the observed.

To compare the co-circulation patterns, we categorized the weeks into 8 possible co-circulation types: 1) none in circulation, i.e., none of the three types/subtypes had ILI+ above their baselines; 2) A(H1N1) alone, i.e., only A(H1N1) had ILI+ above its baseline; and similarly, 3) A(H3N2) alone and 4) B alone; 5) A(H1N1)+A(H3N2), i.e., only A(H1N1) and A(H3N2) had ILI+ above their baselines; 6) A(H1N1)+B, i.e., only A(H1N1) and B had ILI+ above their baselines; 7) A(H3N2)+B, i.e., only A(H3N2) and B had ILI+ above their baselines; and 8) All, i.e., all three types/subtypes had ILI+ above their baselines. We then computed the percentages of weeks falling in each of the 8 co-circulation types and compared them to the observed.

#### 2.2 Results

Figures 8-10 show the model-simulated epidemic trajectories compared to the observations in Hong Kong. As expected, without the constraints by observations (as in a model-inference system), the errors grew quickly and thus the simulated epidemics deviated from the observations. However, overall, simulations with cross-immunity in place were able to better reproduce the observed epidemic pattern (Figs. S8 and S9 vs. Fig. S10). In particular, those simulations were able to reproduce the long periods with near-zero A(H1N1) activities (e.g., 2002-2005) whereas simulations with no cross-immunity (Fig. S10) overestimated the circulation of all types/subtypes, especially for A(H1N1). Comparing the first two scenarios (i.e. both with cross-immunity), simulations with stronger cross-immunity from A(H3N2) against A(H1N1) underestimated the circulation of A(H1N1) and were only able to generate A(H1N1) epidemics following the simulated punctuated antigenic changes when the susceptibility was reset (Fig. S9 vs. Fig. S8 as estimated from the data).

Tally over the entire study period, the first two scenarios (i.e., with cross-immunity) appeared to underestimate the total number of the epidemics (Fig. S11 A and B). However, this was due to the challenge in generating clear bimodal epidemics within a year as those observed in Hong Kong. For instance, there were two consecutive A(H1N1) epidemics in 2008; and similarly, A(H3N2) tended to cause two separate epidemics within a year, e.g., 1999, 2000, 2003, and 2014. The underestimation of number of A(H1N1) epidemics was severer for scenario 2 when stronger cross-immunity from A(H3N2) against A(H1N1) was implemented in the model. Simulations with no cross-immunity were able to generate more epidemics due to overestimation of circulation during periods without observed influenza activity (Fig. S10).

Comparing the 8 co-circulation types (Fig. 11C), scenario 1 using the posterior mean estimates most closely reproduced the observed pattern. In contrast, scenario 2 (i.e., stronger cross-immunity from A(H3N2) against A(H1N1)) largely underestimated the frequency of A(H1N1) epidemics, whereas scenario 3 (i.e., no cross-immunity) largely overestimated co-epidemics caused by multiple types/subtypes.

## Supplementary Table

**Table S1.** Estimated timing of punctuated antigenic changes using a cutoff of 10% increase in susceptibility, instead of 15% in Table 2.

| Strain | Date | SD (days) |
| --- | --- | --- |
| A(H1N1) | 1/21/00 | 4.05 |
|  | 6/11/06 | 0.00 |
|  | 1/25/09 | 0.00 |
|  | 7/6/09 | 3.50 |
|  | 9/13/09 | 2.21 |
|  | 1/4/10 | 6.30 |
|  | 1/14/11 | 3.13 |
| A(H3N2) | 1/18/98 | 0.00 |
|  | 5/15/98 | 6.19 |
|  | 1/10/99 | 4.31 |
|  | 6/13/99 | 12.12 |
|  | 1/9/00 | 1.94 |
|  | 6/27/00 | 5.72 |
|  | 1/13/03 | 9.46 |
|  | 7/11/04 | 0.00 |
|  | 2/25/05 | 14.53 |
|  | 6/20/07 | 3.74 |
|  | 7/28/10 | 3.83 |
|  | 5/10/12 | 8.08 |
|  | 1/2/15 | 4.21 |
|  | 6/6/15 | 8.08 |
| B | 2/6/00 | 0.00 |
|  | 1/28/01 | 9.90 |
|  | 1/27/02 | 0.00 |
|  | 1/6/03 | 7.06 |
|  | 1/13/18 | 0.00 |

## Supplementary figures

**Fig. S1.**
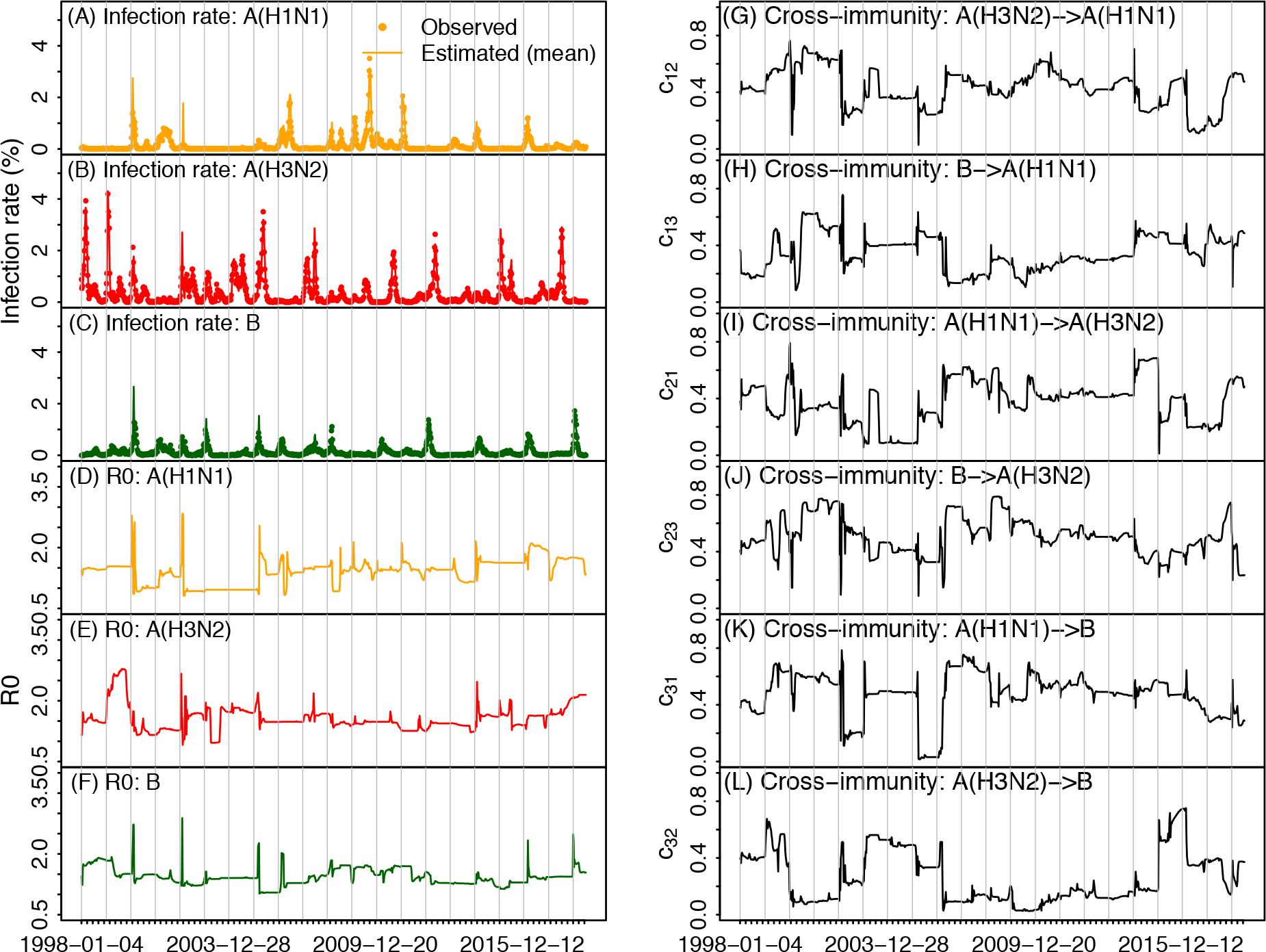
Example model fits and estimates of key parameters using a particle filter. Model fits compared to the observations are shown for A(H1N1) (A), A(H3N2) (B), and B (C). Estimates of *R*_*0*_ are shown in D-F for the three influenza types/subtypes and strength of cross-immunity shown in G-L.

**Fig. S2.**
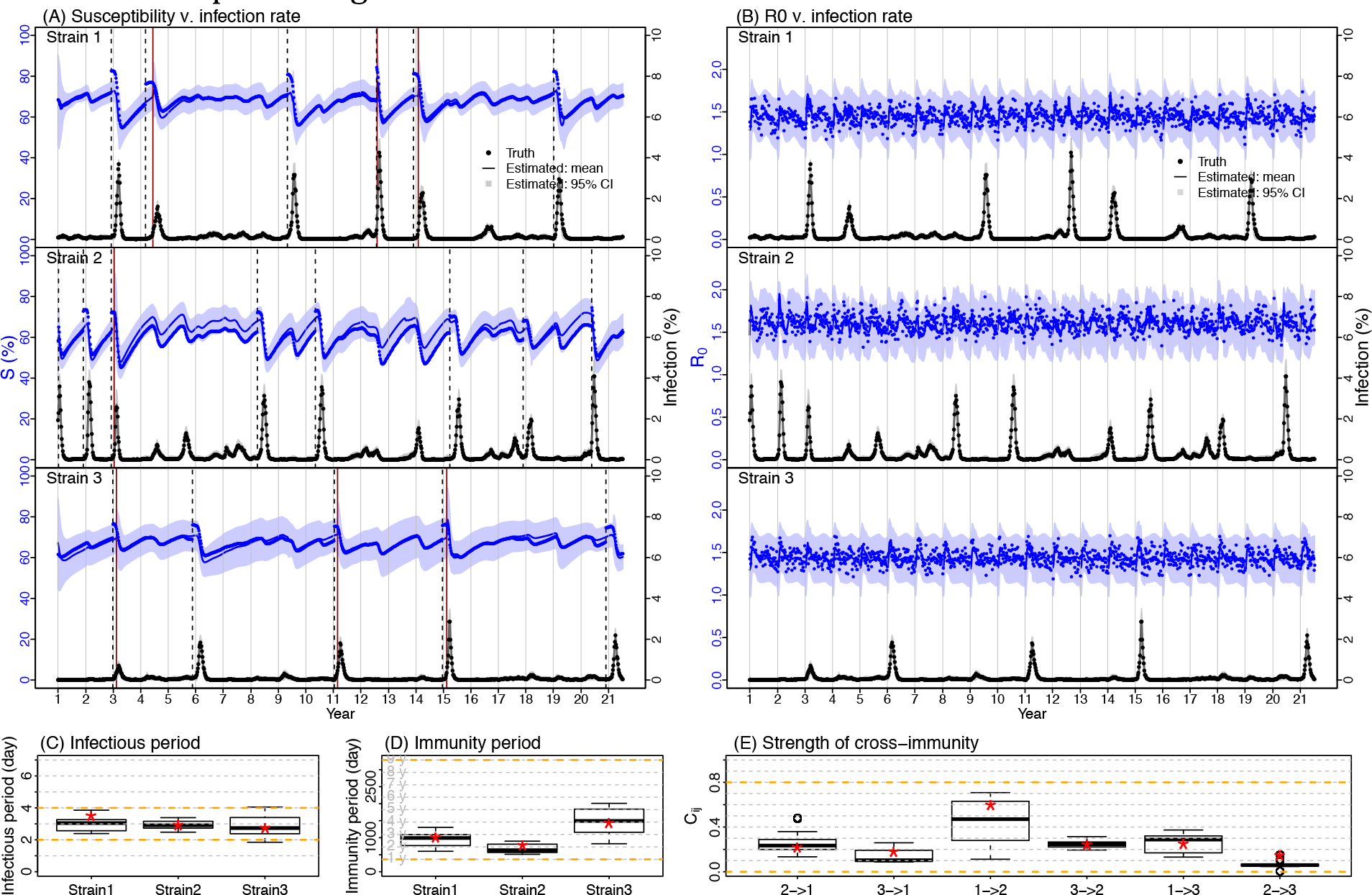
Model validation using the first synthetic dataset. Posterior estimates for the incidence rate and population susceptibility (A), basic reproductive number *R*_*0*_ (B), infectious period (C), immunity period (D), and strength of cross-immunity (E), compared to the true values. In (A) and (B), dots show the true incidence rates (in black; right y-axis), susceptibility (in blue; left y-axis), and *R*_*0*_ (in blue; left y-axis); the lines and surrounding areas show the mean and 95% credible interval (CI) estimates. In (A), vertical dashed black lines show the true weeks with epochal antigenic changes and red lines show model-estimates. In (C)-(E), red stars show true parameter values; box plots show the median, 75%, and 95% CIs of posterior estimates by the model-inference system; orange dashed lines show the prior ranges tested.

**Fig. S3.**
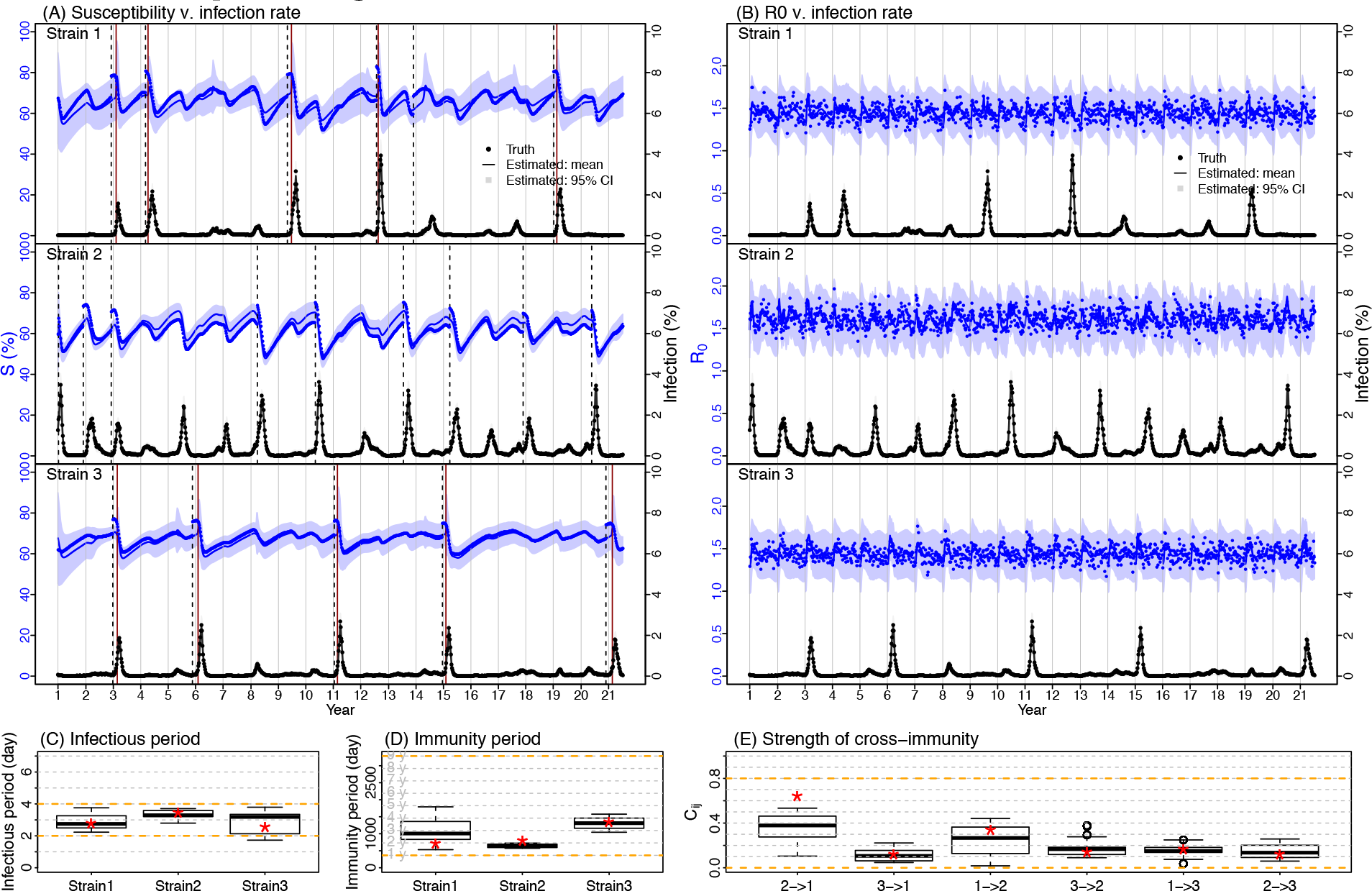
Model validation using the second synthetic dataset. Posterior estimates for the incidence rate and population susceptibility (A), basic reproductive number *R*_*0*_ (B), infectious period (C), immunity period (D), and strength of cross-immunity (E), compared to the true values. In (A) and (B), dots show the true incidence rates (in black; right y-axis), susceptibility (in blue; left y-axis), and *R*_*0*_ (in blue; left y-axis); the lines and surrounding areas show the mean and 95% credible interval (CI) estimates. In (A), vertical dashed black lines show the true weeks with epochal antigenic changes and red lines show model-estimates. In (C)-(E), red stars show true parameter values; box plots show the median, 75%, and 95% CIs of posterior estimates by the model-inference system; orange dashed lines show the prior ranges tested.

**Fig. S4.**
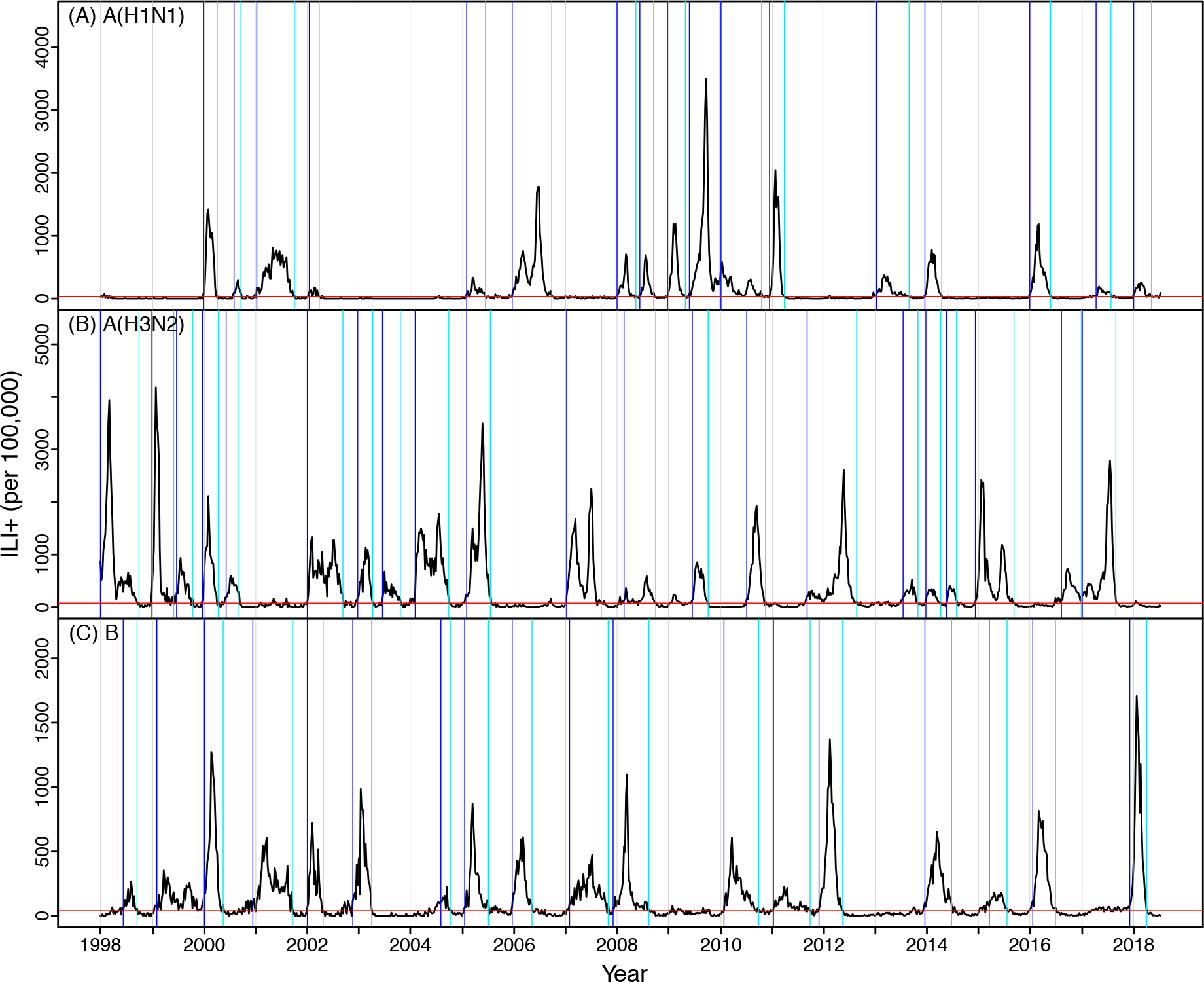
Epidemics for individual types/subtypes as defined by the criteria including small epidemics: (A) A(H1N1), (B) A(H3N2), and (C) B. Black lines are ILI+ observations; red horizontal lines are baselines; blue vertical lines are the identified onsets; cyan vertical lines are identified endings; grey vertical lines are year divisions.

**Fig. S5.**
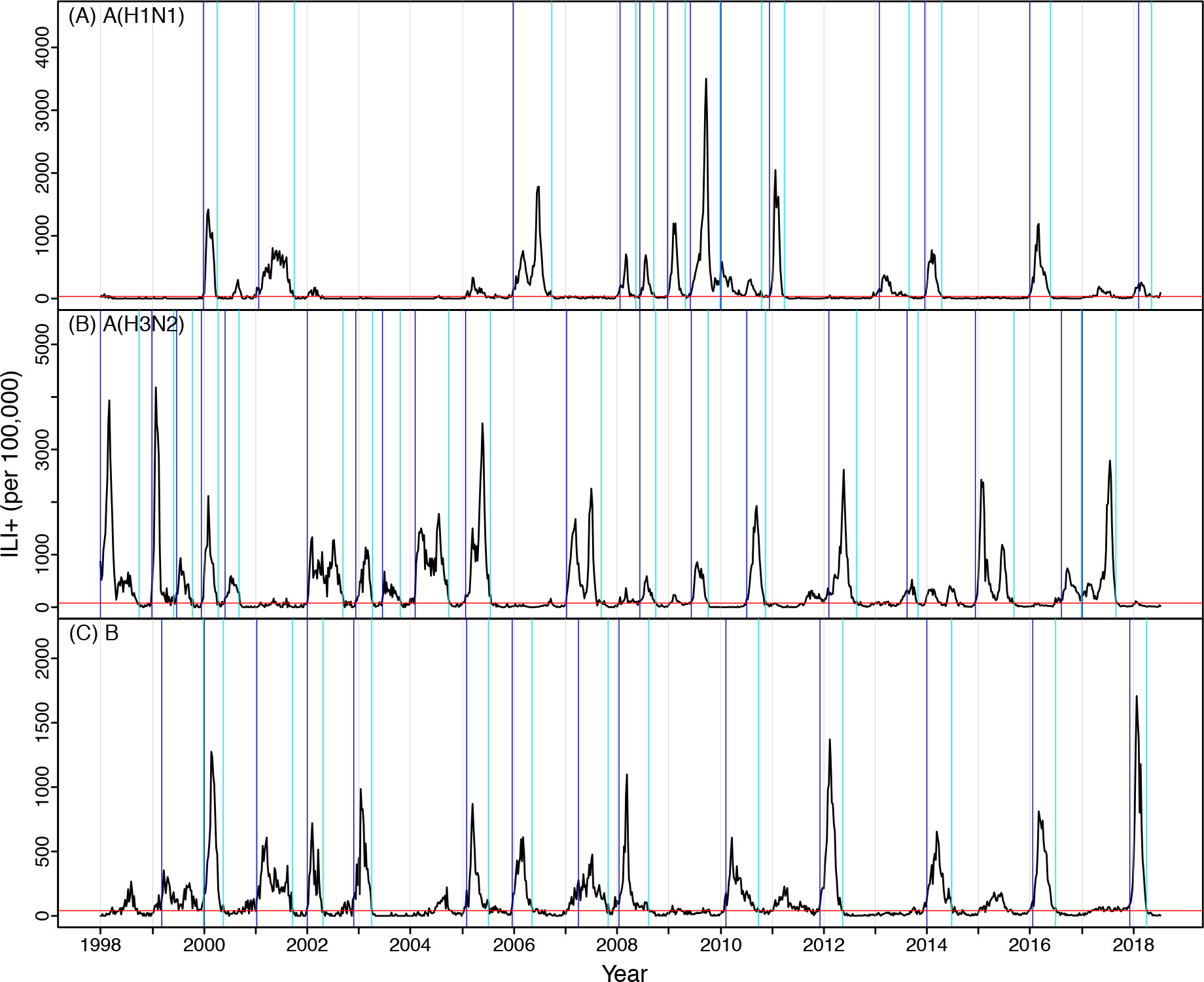
Epidemics for individual types/subtypes as defined by the criteria including only large epidemics: (A) A(H1N1), (B) A(H3N2), and (C) B. Black lines are ILI+ observations; red horizontal lines are baselines; blue vertical lines are the identified onsets; cyan vertical lines are identified endings; grey vertical lines are year divisions.

**Fig. S6.**
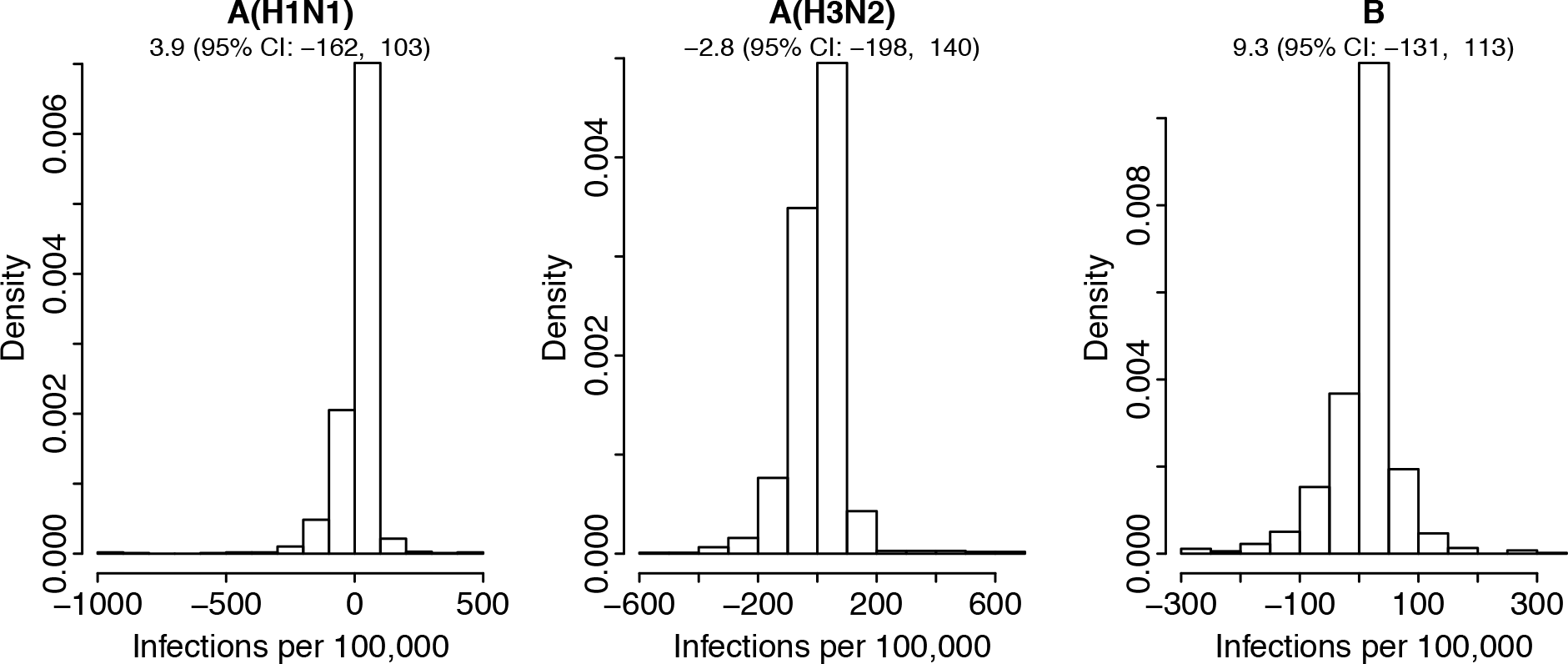
Distributions of residuals of incidence rate estimates. Numbers below the name of virus show the mean and 95% confidence interval in parentheses.

**Fig. S7.**
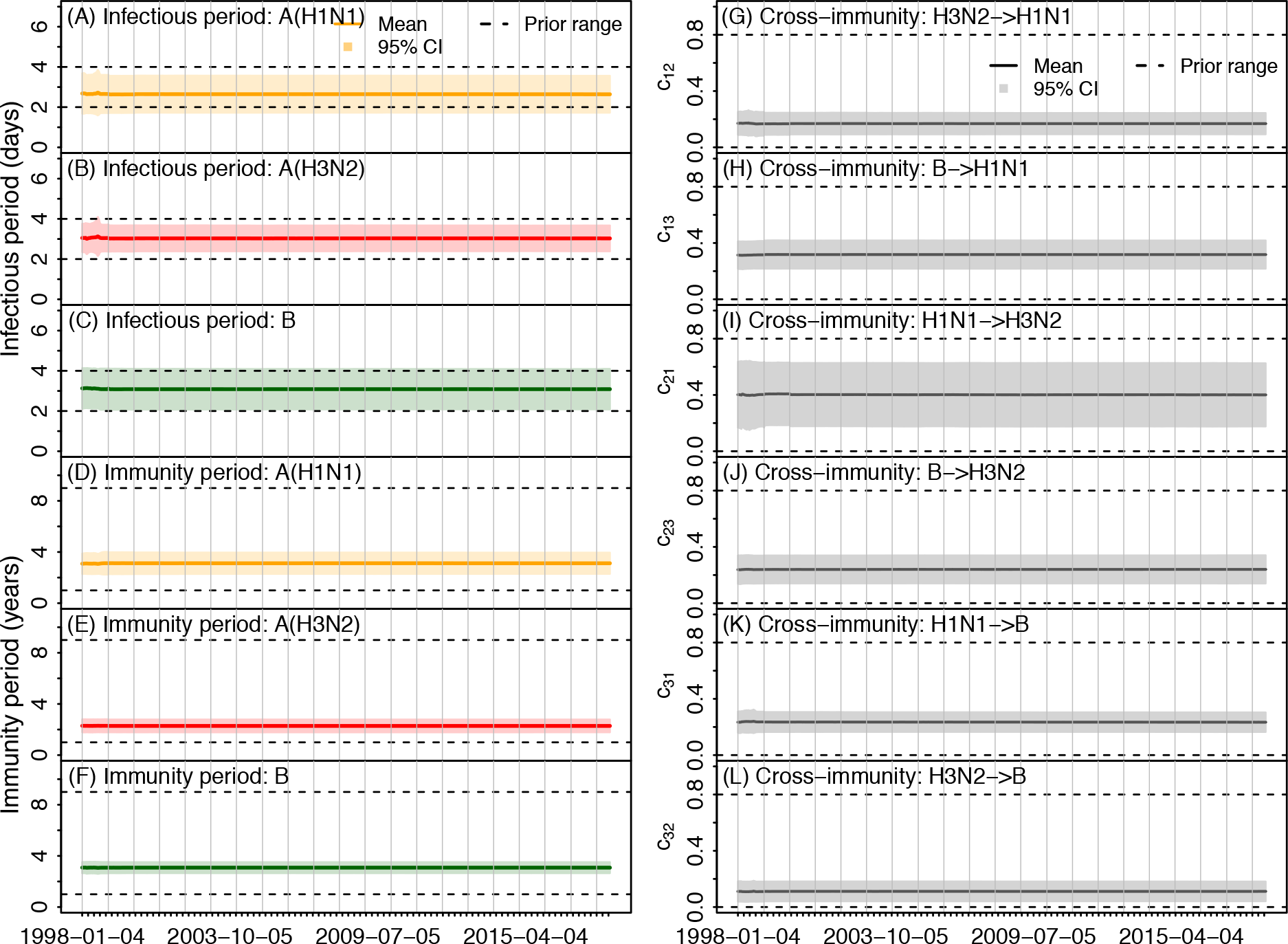
Weekly estimates of key model parameters: infectious period for A(H1N1) (A), A(H3N2) (B), and B (C); immunity period for A(H1N1) (D), A(H3N2) (E), and B (F); and strength of cross-immunity for the six virus-pairs (G-L). Solid lines and surrounding areas show the posterior mean and 95% CI estimates and dashed lines show the prior ranges.

**Fig. S8.**
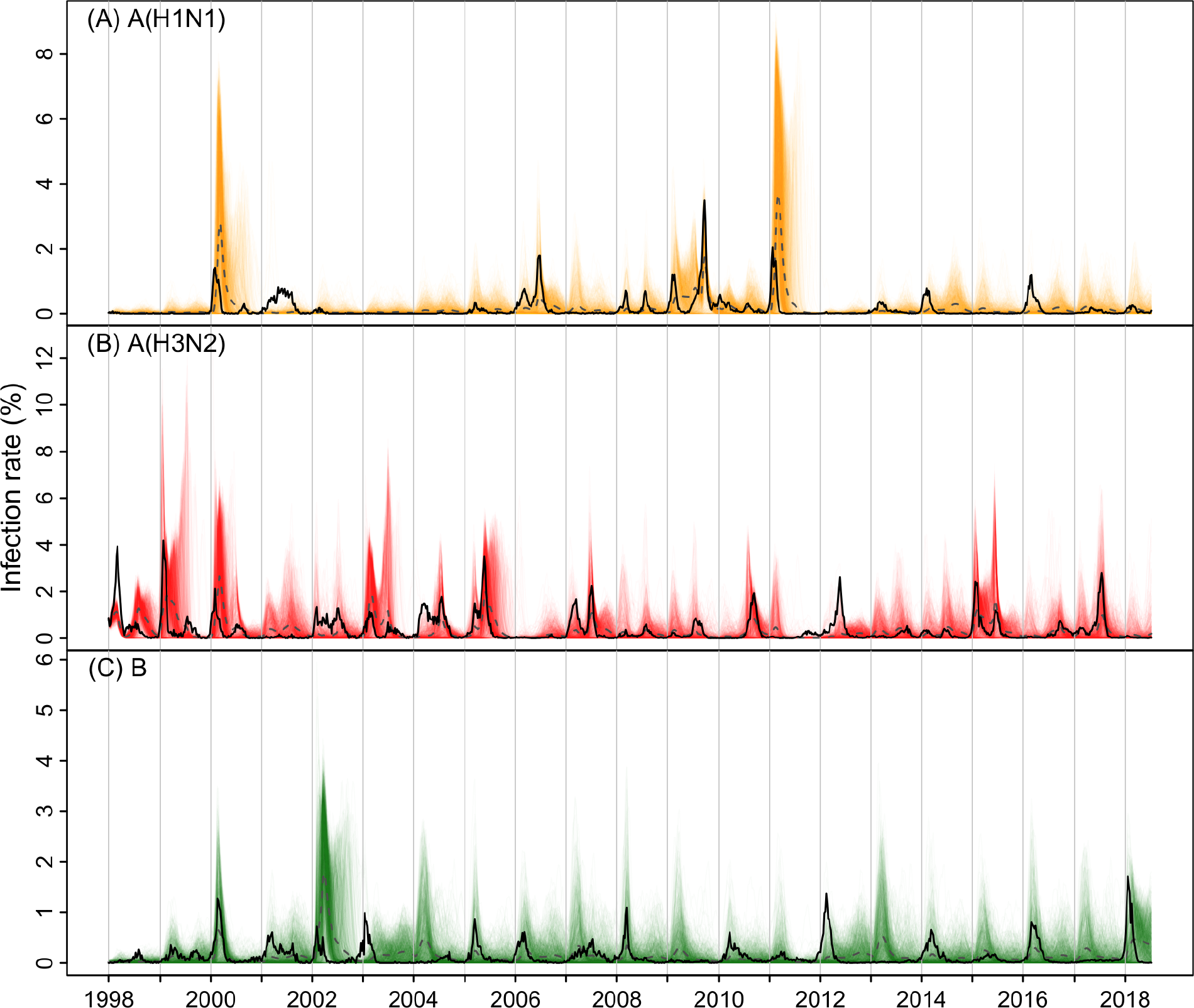
Simulations using the posterior mean estimates. Colored lines show simulated weekly infection rates from 1000 individual stochastic model runs (A(H1N1) in orange, A(H3N2) in red, and B in green); dashed black lines show the weekly mean infection rates across 1000 simulations and solid black lines show the weekly observations for comparison.

**Fig. S9.**
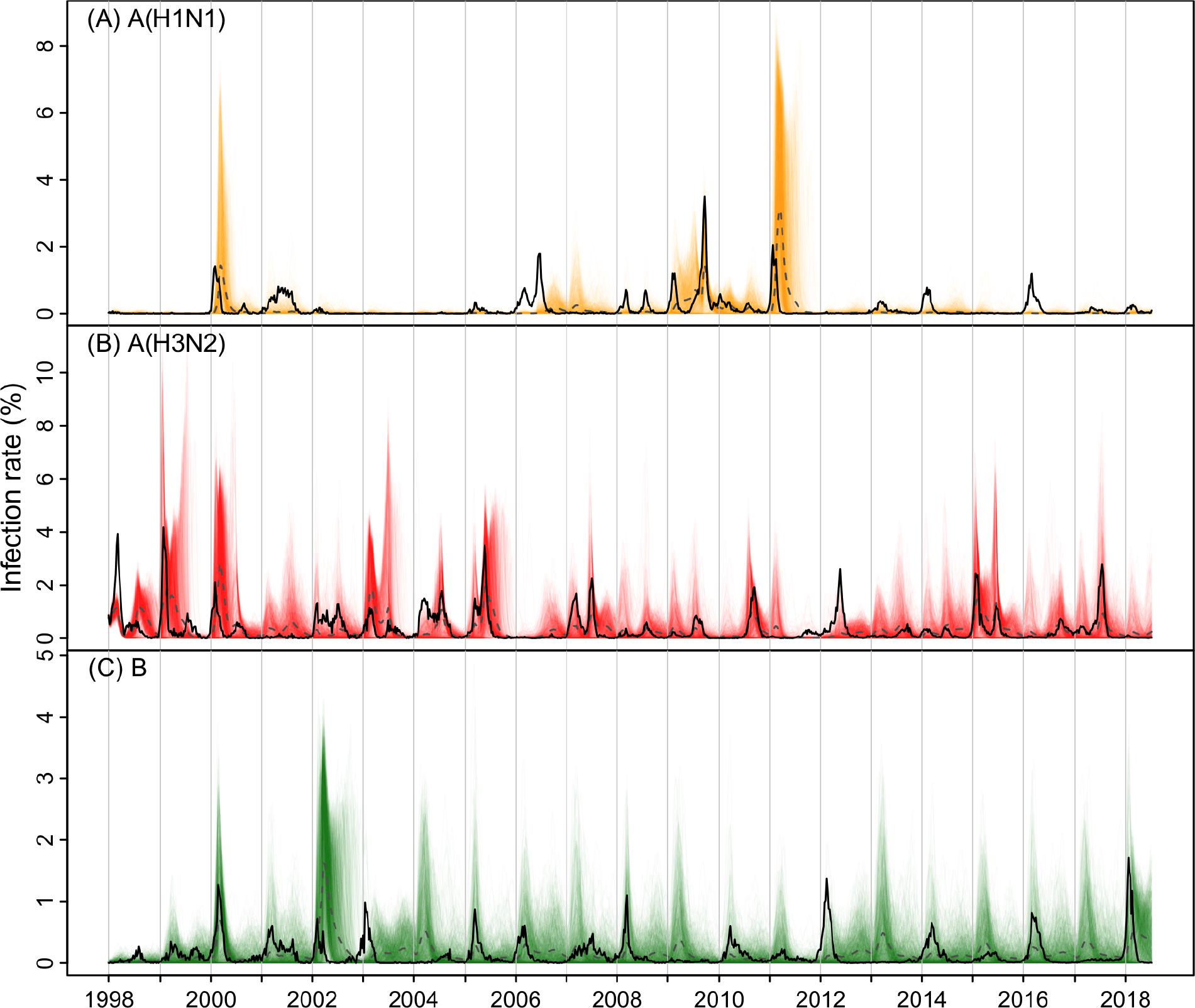
Simulations assuming stronger cross-immunity from A(H3N2) against A(H1N1). The cross-immunity parameter *c*_*H1←H3*_ was set to 0.5 and other parameters set to the posterior mean estimates. Colored lines show simulated weekly infection rates from 1000 individual stochastic model runs (A(H1N1) in orange, A(H3N2) in red, and B in green); dashed black lines show the weekly mean infection rates across 1000 simulations and solid black lines show the weekly observations for comparison.

**Fig. S10.**
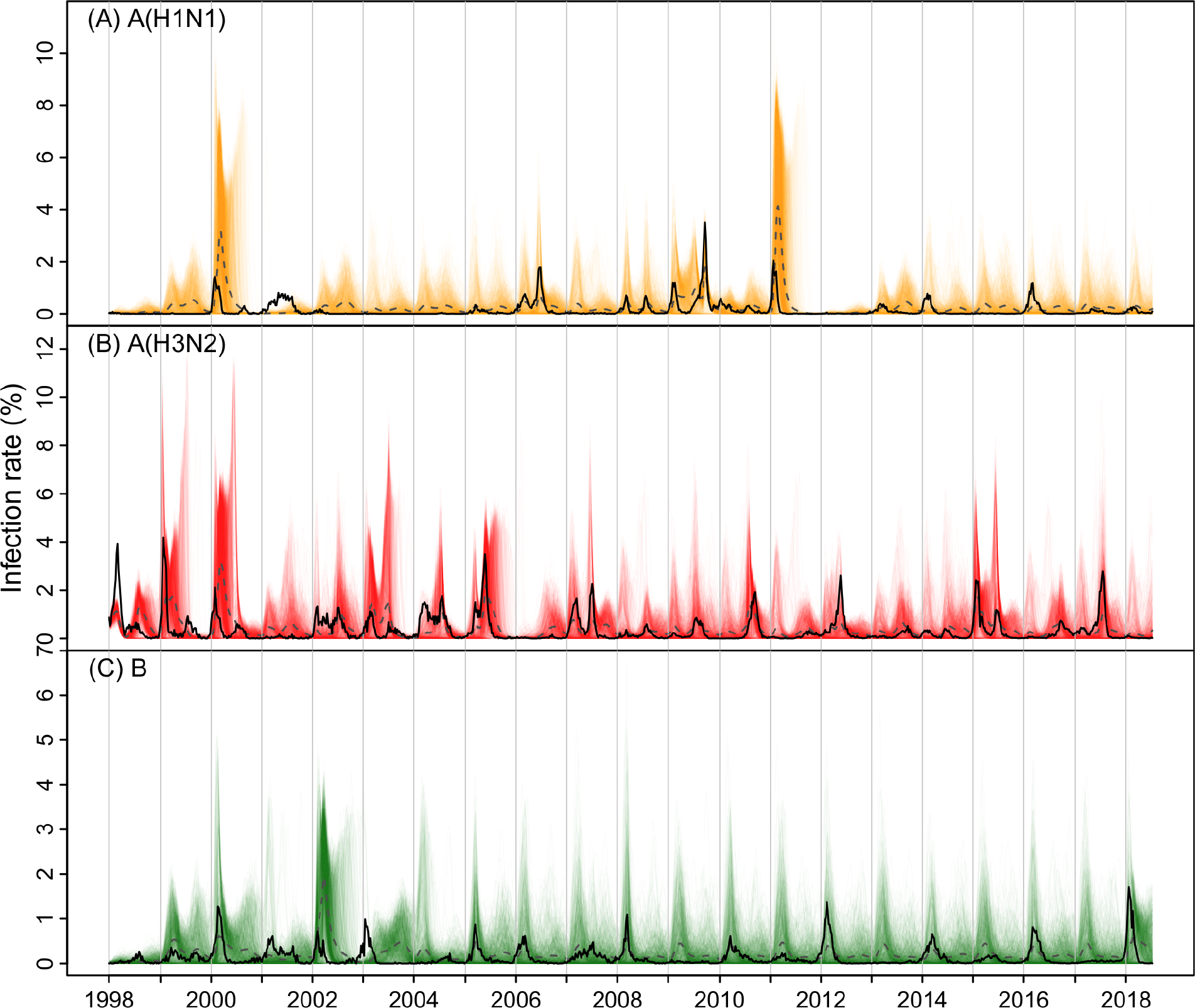
Simulations assuming no cross-immunity. All cross-immunity terms were set to 0 and other parameters set to the posterior mean estimates. Colored lines show simulated weekly infection rates from 1000 individual stochastic model runs (A(H1N1) in orange, A(H3N2) in red, and B in green); dashed black lines show the weekly mean infection rates across 1000 simulations and solid black lines show the weekly observations for comparison.

**Fig. S11.**
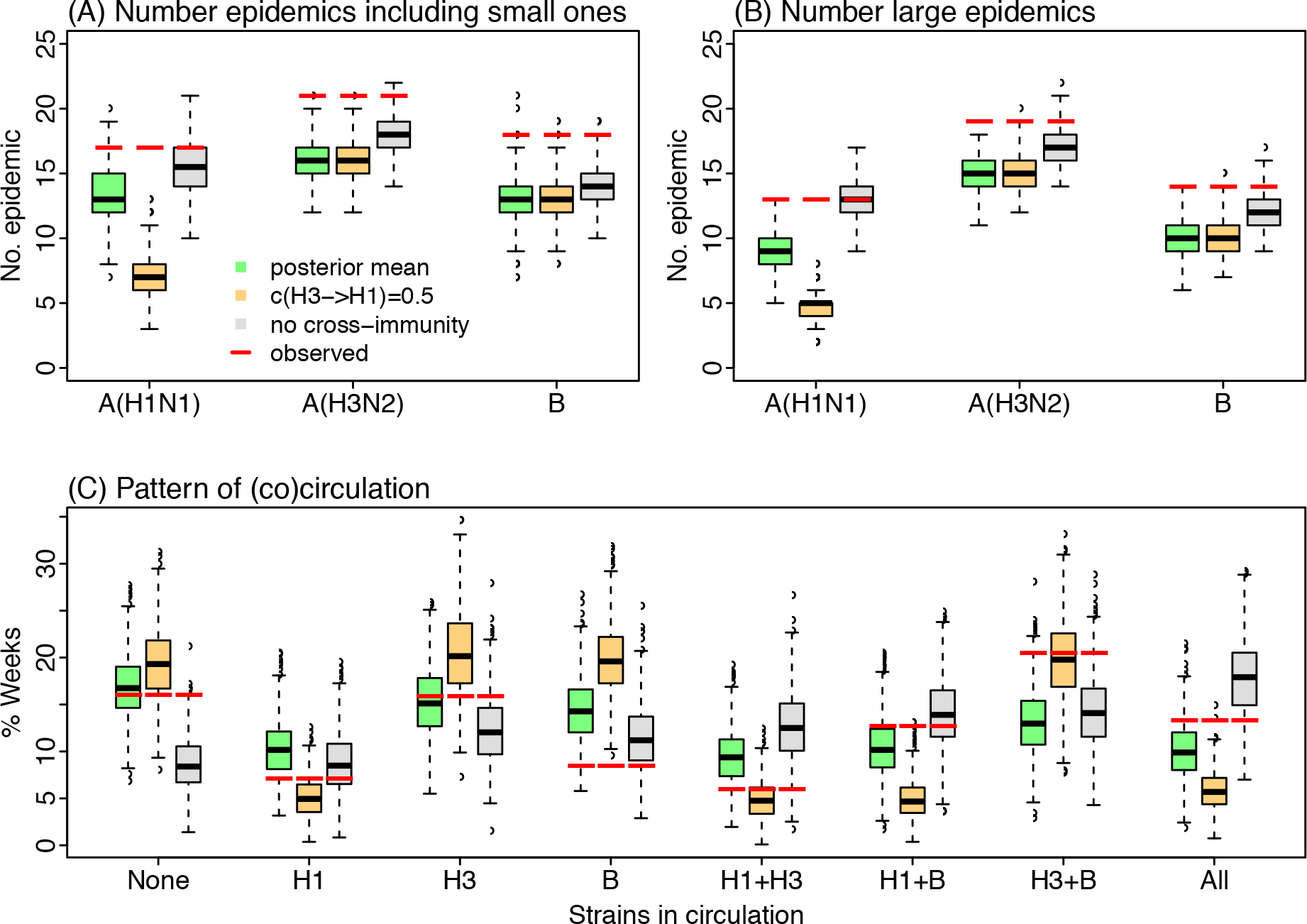
Epidemic and co-circulation patterns under different strengths of cross-immunity. Box and whisker plots show the distributions across 1000 simulations for each cross-immunity scenario: scenario 1 as estimated in green; scenario 2 with stronger cross-immunity from A(H3N2) against A(H1N1) in orange; and scenario 3 with no cross-immunity in grey. Horizontal thick black lines show the median; box edges show the 25^th^ and 75^th^ percentiles; the whiskers show the full ranges and dots show outliers. Red segments show the corresponding observations in Hong Kong.

